# Report 32: Age groups that sustain resurging COVID-19 epidemics in the United States

**DOI:** 10.1101/2020.09.18.20197376

**Authors:** Mélodie Monod, Alexandra Blenkinsop, Xiaoyue Xi, Daniel Hebert, Sivan Bershan, Simon Tietze, Valerie C Bradley, Yu Chen, Helen Coupland, Sarah Filippi, Jonathan Ish-Horowicz, Martin McManus, Thomas Mellan, Axel Gandy, Michael Hutchinson, H Juliette T Unwin, Michaela A C Vollmer, Sebastian Weber, Harrison Zhu, Anne Bezancon, Neil M Ferguson, Swapnil Mishra, Seth Flaxman, Samir Bhatt, Oliver Ratmann, on behalf of the Imperial College COVID-19 Response Team

## Abstract

Following initial declines, in mid 2020, a resurgence in transmission of novel coronavirus disease (COVID-19) has occurred in the United States and parts of Europe. Despite the wide implementation of non-pharmaceutical interventions, it is still not known how they are impacted by changing contact patterns, age and other demographics. As COVID-19 disease control becomes more localised, understanding the age demographics driving transmission and how these impacts the loosening of interventions such as school reopening is crucial. Considering dynamics for the United States, we analyse aggregated, age-specific mobility trends from more than 10 million individuals and link these mechanistically to age-specific COVID-19 mortality data. In contrast to previous approaches, we link mobility to mortality via age specific contact patterns and use this rich relationship to reconstruct accurate transmission dynamics. Contrary to anecdotal evidence, we find little support for age-shifts in contact and transmission dynamics over time. We estimate that, until August, 63.4% [60.9%-65.5%] of SARS-CoV-2 infections in the United States originated from adults aged 20-49, while 1.2% [0.8%-1.8%] originated from children aged 0-9. In areas with continued, community-wide transmission, our transmission model predicts that re-opening kindergartens and elementary schools could facilitate spread and lead to additional COVID-19 attributable deaths over a 90-day period. These findings indicate that targeting interventions to adults aged 20-49 are an important consideration in halting resurgent epidemics and preventing COVID-19-attributable deaths when kindergartens and elementary schools reopen.

**One sentence summary:** Adults aged 20-49 are a main driver of the COVID-19 epidemic in the United States; yet, in areas with resurging epidemics, opening schools will lead to more COVID-19-attributable deaths, so more targeted interventions in the 20-49 age group could bring epidemics under control, avert deaths, and facilitate the safe reopening of schools.

## 1 Introduction

In 2020 a novel pathogen, severe acute respiratory syndrome coronavirus 2 (SARS-CoV-2) emerged in Hubei Province, China [1]. Spread within China occurred in January 2020 and the resultant disease was named COVID-19. Following worldwide spread, the implementation of large-scale non-pharmaceutical interventions has led to sustained declines in the number of reported SARS-CoV-2 infections and deaths. However since mid June, the daily number of reported COVID-19 cases has re-surged in the United States, surpassing 40,000 daily reported cases on June 26 [2], and increasing daily cases are beginning to be reported in Europe [3]. Demographic analyses of reported cases have suggested that individuals aged 20 − 49 may be driving the re-surging epidemic [4, 5]. Here, we use detailed, longitudinal, and age-specific population mobility and COVID-19 mortality data to estimate how non-pharmaceutical interventions, changing contact intensities interplay, age and other factors have led to resurgent disease spread. We identify the population age groups driving SARS-CoV-2 spread in 35 U.S. states, the District of Columbia and New York City through August 23, 2020, and quantify the likely impact of school reopening on case and death counts under the scenario that transmission from the age groups that primarily drive transmission continues uninterrupted.

Similar to many other respiratory diseases, the spread of SARS-CoV-2 occurs primarily through close human contact, which, at a population level, is highly structured [6]. Prior to the implementation of COVID-19 interventions, contacts concentrated among individuals of similar age, were highest among school-aged children, and also common between children and their parents, and middle-aged adults and the elderly [7]. Since the beginning of the pandemic, these contact patterns have changed substantially [8, 9, 10]. In the United States, the Berkeley Interpersonal Contact Study suggests that in late March 2020 after stay-at-home orders were issued, the average number of daily contacts made by a single individual, also known as contact intensity, dropped to four or fewer contacts per day [10]. Data from China indicate that infants and school-aged children had almost no contact to similarly aged children in the first weeks after stay-at-home orders, and reduced contact intensities with older individuals [8]. However, detailed age-specific population-level contact and mobility data have remained scarce, especially longitudinally, and this has impeded a better understanding of the age-specific sources driving COVID-19 transmission.

## 2 Results

### Fine scale mobility trends across the United States

We compiled a national-level, aggregate mobility data set using cell phone data from >10 million individuals with Foursquare’s location technology, Pilgrim [11], which leverages a wide variety of mobile device signals to pin-point the time, duration, and location of user visits to locations such as shops, parks, or universities. Unlike the population-level mobility trends published by Google from cell phone geolocation data [12], the data are disaggregated by age. User visits were analyzed from February 1, 2020, aggregated, and projected to estimate for each state and two metropolitan areas daily foot traffic for individuals aged 18 − 24, 25 − 34, 35 − 44, 45 − 54, 55 − 64, and 65+ years. To obtain age-specific mobility trends, the data were divided by the corresponding averages in the baseline period February 3 - February 9, 2020 per age band and state or metropolitan area (see Supplementary Material S1).

Across the US as a whole, the mobility trends indicate substantial initial declines in extra-household visits (location an individual spends time at that is not the primary residence) followed by a subsequent rebound for all age groups (Figure 1A; see also Supplementary Figure S12). During the initial phase of the epidemic, trends declined most strongly among individuals aged 18-24 years across almost all states and metropolitan areas, and subsequently tended to increase most strongly among individuals aged 18-24 in the majority of states and metropolitan areas (Supplementary Figure S1), consistent with re-opening policies for restaurants, night clubs, and other venues [13]. Yet, by the last observation week August 15, 2020 - August 21, 2020, the data suggest mobility levels continue to be below those observed in the baseline period February 3 to February 9, 2020, in most states and metropolitan areas (Figure 1B). In addition, considering both the initial decline and subsequent rebound, our data indicate that mobility levels among individuals aged < 35 years have not increased significantly above those observed among individuals aged 35-44, and that as of August 2020 there have been no significant shifts in the relative levels of mobility between age groups (Figure 1A-B, and Supplementary Figure S13).

**Figure 1:**
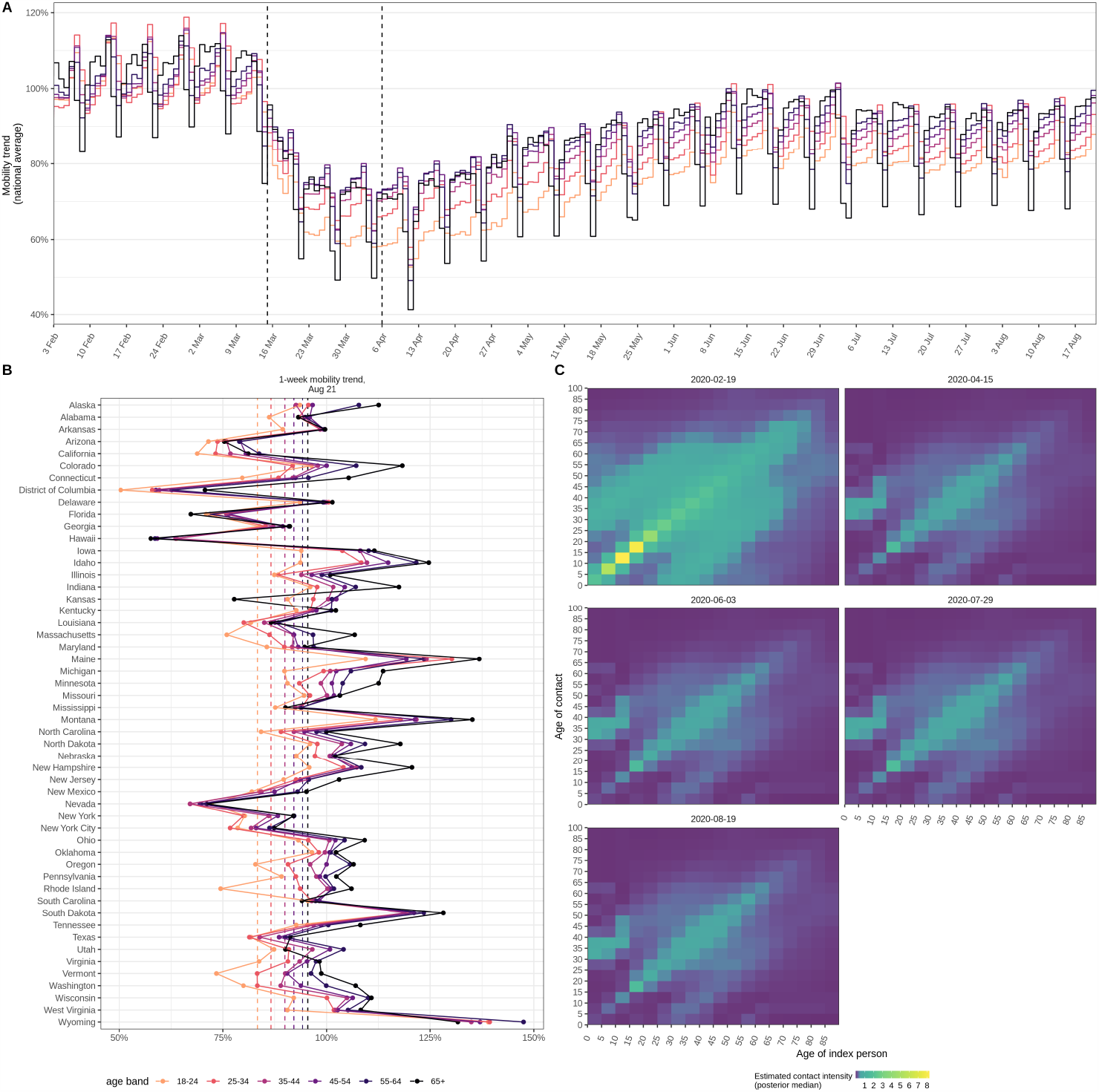
Mobility trends, and estimated time evolution of contact intensities in the United States. (**A**) National, longitudinal mobility trends for individuals aged 18-24, 25-34, 35-44, 45-54, 55-64, 65+, relative to the baseline period February 3 to February 9, 2020. The vertical dashed lines show the dip and rebound dates since when mobility trends began to decrease and increase, which were estimated from the time series data. (**B**) 1-week average of age-specific mobility trends between August 15, 2020 - August 21, 2020 across the United States. (**C**) Inferred time evolution of contact intensities in California.

Mobile phone signals are challenging to analyse, owing e.g. to daily fluctuations in the user panel providing location data, imprecise geolocation measurements, and changing user behaviour [14]. We cross-validated the inferred mobility trends against age-specific mobility data from a second mobile phone intelligence provider, Emodo. This second data set also showed no evidence for significant shifts in relative mobility levels between age groups (see Supplementary Material S1), leading us to hypothesize that the resurgent epidemics in the United States may not be a result of changes in the contribution of different age groups to SARS-CoV-2 transmission.

### Bayesian semi-mechanistic contact and infection model to characterise age-specific SARS-Cov-2 transmission

To test this hypothesis, we incorporated the mobility data into a Bayesian contact-and-infection model that describes time-changing contact and transmission dynamics at state and metropolitan area-level across the United States (see Supplementary Figure S2 and Supplementary Text S3). For the time period prior to changes in mobility trends, we used data from pre-COVID-19 contact surveys [6], and each state or metropolitan area’s age composition and population density to predict contact intensities between individuals grouped in 5-year age bands. On weekends, contact intensities between school-aged children are lower than on weekdays, while inter-generational contact intensities are higher. In the model, the observed age-specific mobility trends of Figure 1 are then used to estimate in each location (state or metropolitan area) daily changes in age-specific contact intensities for individuals aged 15 and above. We assumed that the effect of the observed mobility trends on changing contact intensities was the same across age groups. For younger individuals, for who mobility trends are not recorded, contact intensities during school closure periods were set to estimates from two contact surveys conducted post lockdown [9, 8]. In turn, the contact intensities are used to estimate the rate of SARS-CoV-2 transmission, and subsequently infections and deaths.

An important feature of SARS-CoV-2 transmission is that similarly to other coronaviruses but unlike pandemic influenza [15], susceptibility to SARS-CoV-2 infection increases with age [8, 16, 17]. Here, we used contact tracing data from Hunan province, China [8] to specify lower susceptibility to SARS-CoV-2 infection among children aged 0-9, and higher susceptibility among individuals aged 60+, when compared to the 10-59 age group. Previously infected individuals are assumed to be immune to re-infection within the 6-month analysis period, consistent with mounting evidence for sustained antibody responses to SARS-CoV-2 antigens [18].

In the United States, COVID-19 epidemic trajectories differ substantially across locations and over time, and apart from mobility trends, other factors such as adherence to social distancing guidelines and consistent face mask use contribute to the extent to which spread of SARS-CoV-2 is limited [19, 20]. Thus, and following earlier work [21], the model incorporates random effects in space and time to allow for unobserved factors that could modulate disease-relevant behaviour and contact patterns.

### Age groups sustaining SARS-CoV-2 spread in the United States

To disentangle the contribution of different age groups to onward infection, we recorded age-specific, COVID-19-attributed mortality data from 40 U.S. states, the District of Columbia and New York City since March 15, 2020 (Supplementary Text S2 and [22]). Then, we fitted the contact-and-infection model in a Bayesian framework to the mobility trends and the mortality time series data from 35 U.S. states, the District of Columbia and New York City with at least 300 COVID-19-attributed deaths. Kansas was excluded due to atypical mobility trend data, giving a total of 5,579 observation days. The estimated disease dynamics closely reproduced the age-specific COVID-19 death counts (Supplementary Figure S3).

Figure 2 illustrates the model fits for New York City, Florida, California, and Arizona, showing that the inferred epidemic dynamics differed markedly across states and metropolitan areas. In New York City, the epidemic accelerated for at least 4 weeks since the 10th cumulative death and until age-specific reproduction numbers started to decline, resulting in an epidemic of large magnitude as shown through the estimated number of infectious individuals (Figure 2, mid column). Subsequently, we find that reproduction numbers for all age groups were controlled to below one except a two-week period in June (Figure 2, rightmost column), resulting in a steady decline of infectious individuals. In Florida, we estimate reproduction numbers remained above one for individuals aged 20-49, and in June increased substantially above one for individuals aged 10-64, resulting in a moderate initial decline in infectious individuals followed by a peak in the number of infectious individuals in late July, and subsequent decline. In California, we estimate that reproduction numbers for individuals aged 35-49 remained above one throughout the pandemic, and in June increased to above one for individuals aged 20-64, resulting in a similar but less marked increase in infectious individuals when compared to Florida. In Arizona, we estimate reproduction numbers remained above one for individuals aged 10-49, and fell below one in August, resulting in a sustained increase in infectious individuals until August, and subsequent decline. More detailed situation analyses for all locations are presented in Supplementary Text S7.

**Figure 2:**
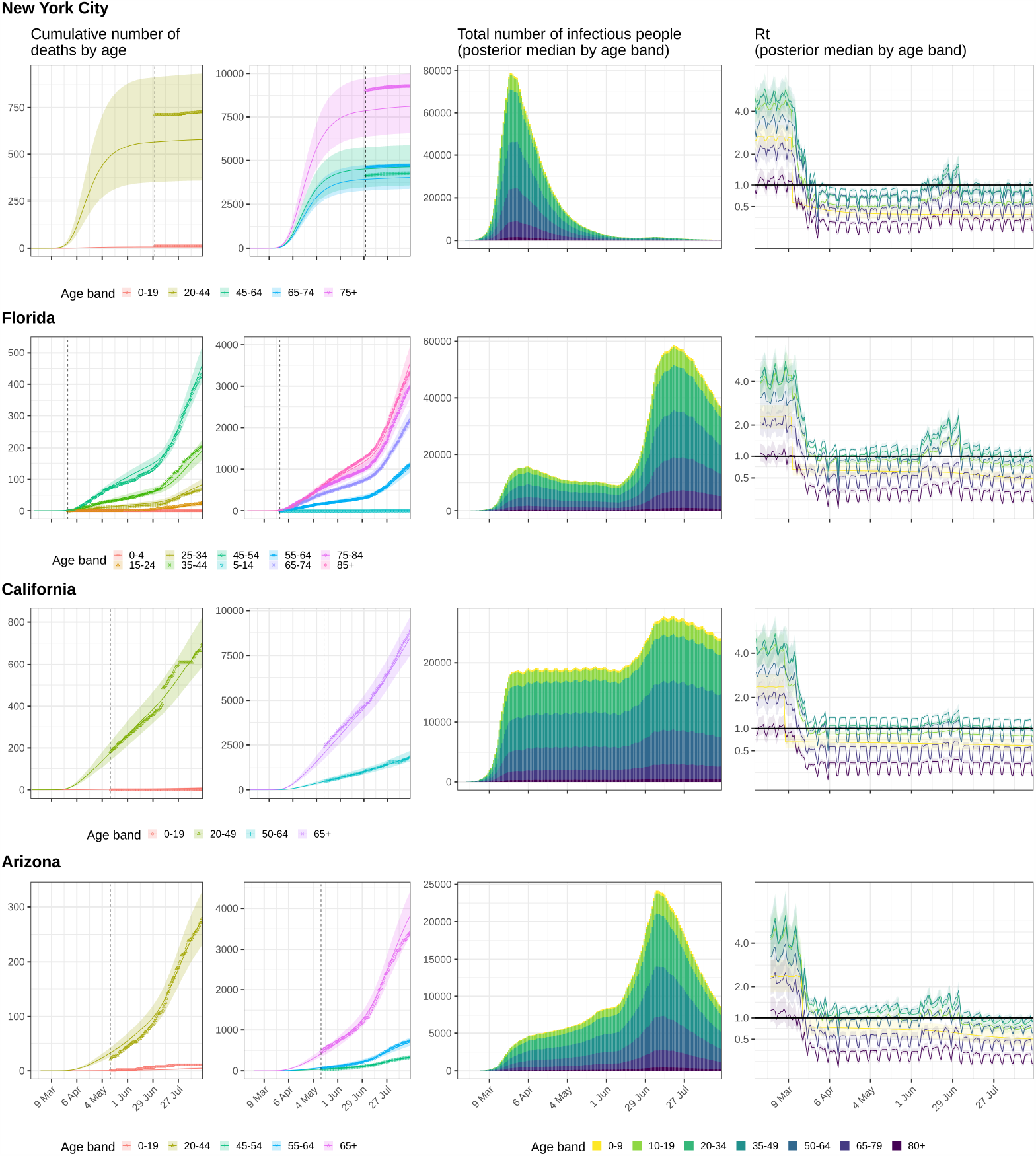
Model fits and key generated quantities for New York City, California, Florida and Arizona. (**left**) Observed cumulative COVID-19 mortality data (dots) versus posterior median estimates (line) and 95% credible intervals (ribbon). The vertical line indicates the collection start date of age-specific death counts. (**middle**) Estimated number of infectious individuals by age (posterior median). (**right**) Estimated age-specific effective reproduction number, posterior median (line) and 95% credible intervals (ribbon).

Figure 3 summarises the epidemic situation for all states and metropolitan areas evaluated. Children aged 0-9 and adults aged 65+ consistently had the lowest estimated reproduction numbers, and these typically remained below one since mobility trends began to decline in March 2020 (Supplementary Table S1), which is consistent with the low contact intensities from these age groups during school closure periods. By August 17, 2020, the estimated reproduction number across all locations evaluated was above one only for individuals aged 35-49 (1.10 [1.04-1.17]), and close to one for individuals aged 10 − 19 or 20 − 34 (Supplementary Table S2). This suggests that targeted interventions to these age groups, and in particular adults aged 35-49, could bring resurgent COVID-19 epidemics under control.

**Figure 3:**
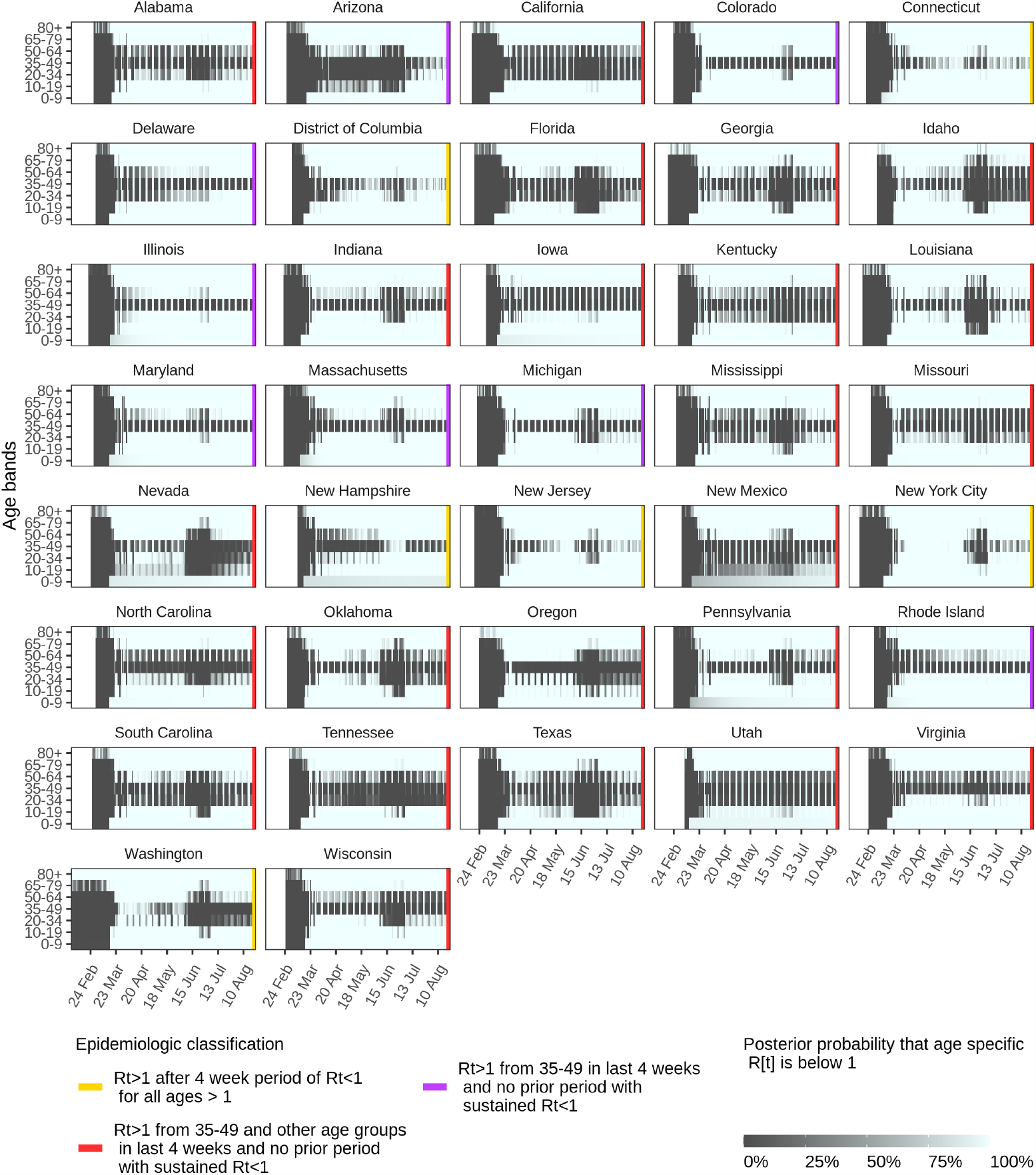
Time evolution of estimated age-specific SARS-CoV-2 transmission dynamics in the United States. Each panel shows for the corresponding location (state or metropolitan area) the estimated posterior probability that the daily effective reproduction number from individuals stratified in 7 age groups were below. Darker colours indicate low probability that reproduction numbers were below one. Colour codes on the right associated each location to one five characteristic patterns of disease spread in the United States.

To quantify the contribution of each age group to onward transmission, we also considered the reconstructed transmission flows, because reproduction numbers estimate the number of secondary infections per infected individual, and the number of infectious individuals varies by age as a result of age-specific susceptibility gradients and age-specific contact exposures. Cumulating over time and across all locations evaluated, we estimate that the percent contribution to onward spread was 35.4% [34.2%-36.5%] from individuals aged 35-49, compared to 1.3% [0.8%-2.0%] from individuals aged 0-9, 10.1% [9.2%-11.0%] from individuals aged 10-19, 28.3% [26.9%-29.5%] from individuals aged 20-34, 18.6% [18.1%-19.2%] from individuals aged 50-64, 5.5% [3.7%-8.1%] from individuals aged 65-79 age group, and 0.6% [0.4%-0.9%] from individuals aged 80+ (Table 1). Supplementary Figure S4 compares the contributions of each age group to SARS-Cov-2 transmission against the population age composition in each state. Over time, the model estimates that the mean age of new SARS-CoV-2 infections has been remarkably constant, showing that shifts in age-specific transmission dynamics are not required to explain heterogeneous and resurgent disease dynamics across the United States (Supplementary Figures S5 and S6).

**Table 1:**
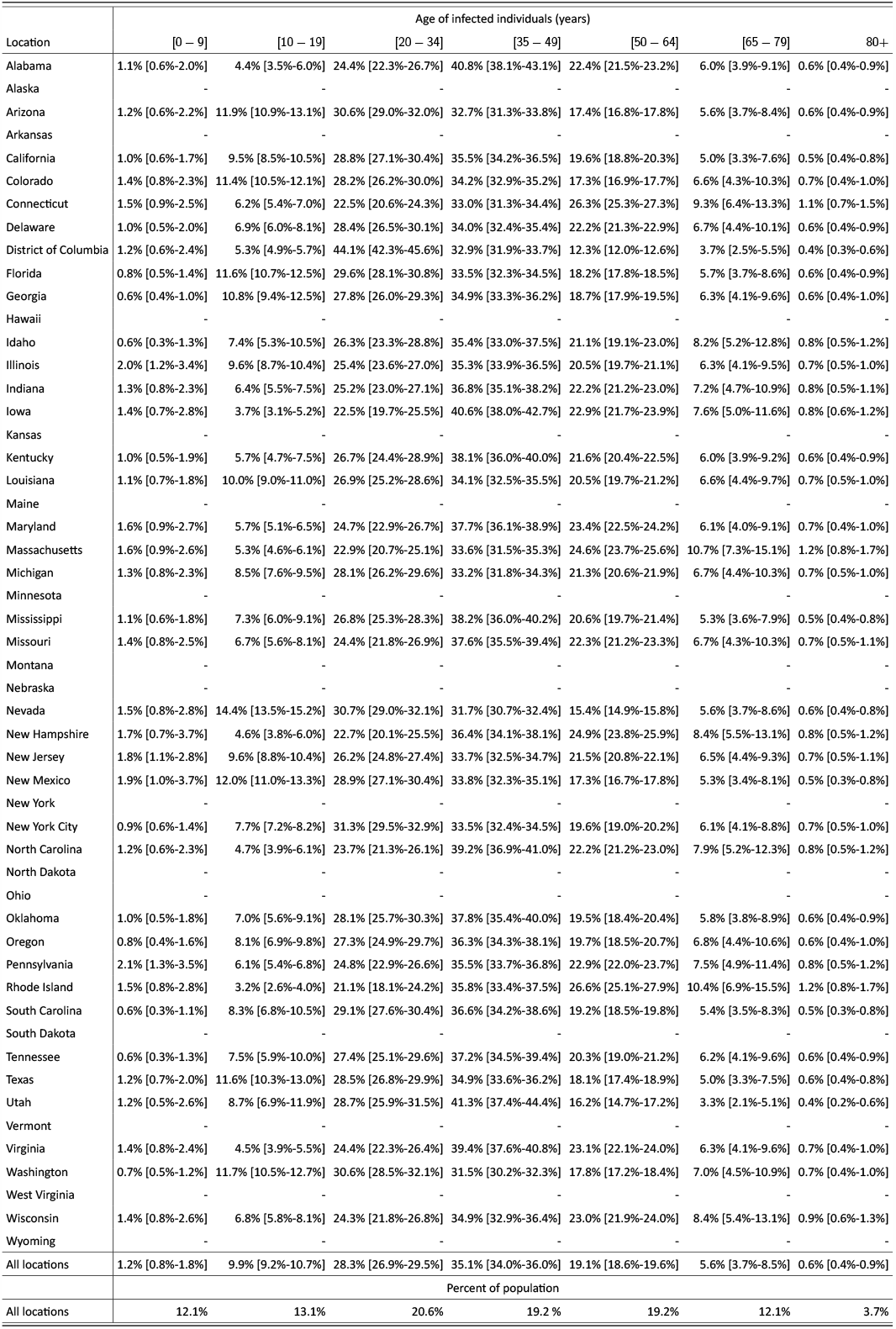
**Estimated cumulated contribution of age groups to SARS-CoV-2 transmission until August 17, 2020**. Posterior median estimates for the United States, and each location (state or metropolitan area), along with 95% credible intervals.

### School opening scenarios

The epidemic situation could change as schools re-open across the United States in August and September, 2020, especially in areas with resurgent, community-wide transmission primarily from adults. Re-opening kindergartens and elementary schools for children aged 0-11 are a national priority [23]. We thus focused on school opening scenarios in which children aged 0-11 return to engage in typical contact patterns with their peers and older individuals, while mobility levels, reproduction numbers, and the transmission potential of all other age groups were kept fixed as inferred by the end of August 2020 for the forecast period. We assumed disease transmission from and to children aged 0-11 is reduced by 50% due to face mask use and other non-pharmaceutical interventions [19], considering also the range 0%-80%. The scenarios were evaluated over 90 days and contrasted to continued school closure scenarios. Across all 37 states and metropolitan areas evaluated, we estimate by November 24, 2020 a 253.7% [199.3%-366.9%] increase in infections among children aged 0-11, and 24 [13, 42] excess COVID-19 attributable deaths among children aged 0-11, resulting in pediatric COVID-19 attributable mortality figures that are similar to pediatric influenza-like mortality (Table S4). The forecasts further estimate 6,181 [3,286, 11,925] excess COVID-19 attributable deaths in the total population, which is a 12.6% [7.4%-22.7%] increase compared to the continued school closure scenario, by November 24, 2020 (Table S4). In the central analysis, the predicted excess COVID-19 attributable deaths are concentrated in areas with resurgent epidemics, most notably Texas, California and Florida, and few additional COVID-19 attributable deaths are predicted in areas where reproduction numbers from individuals aged 20 − 49 are below one or close to one (Figure S7). We emphasise that the predictions depend on the assumed level of transmission reductions in kindergartens and elementary schools, with no substantial increases in COVID-19 deaths when transmission from and to children aged 0-11 is reduced by 66% or more, and substantial increases in COVID-19 attributable deaths in most states and metropolitan areas when transmission from and to children aged 0-11 is not reduced due to pre-cautionary measures (Figures S8-S10, Tables S5-S7).

### Limitations

The findings of this study need to be considered in the context of the following limitations. First, we rely on limited data from two contact surveys performed in the United Kingdom and China to characterise contact patterns from and to younger individuals during school closure periods [9, 8]. We explored the impact of higher inter-generational contact intensities involving children during school closure periods, and in these analyses the estimated contribution of children aged 0-9 to onward spread until August 2020 remained below 2% (Supplementary Material S6). Second, while COVID-19 deaths are considered a more robust measure of SARS-CoV-2 spread than reported cases due to the high proportion of asymptomatic cases [24], epidemiologic models are sensitive to assumptions on the infection fatality ratio (IFR) that relates infections to deaths. We reconsidered a recent meta-analysis of estimates from large-scale seroprevalence studies [25], and found greater uncertainty associated with IFR estimates for individuals below age 40 (Supplementary Text S3). Using these uncertainty ranges in the model, we estimate greater COVID-19 burden among individuals below age 40, and we are able to match data from several sero-prevalence surveys conducted by the Centers for Disease Control and Prevention [26] (Supplementary Table S3 and Supplementary Text S5). Third, we cannot rule out that the observed time evolution of age-specific COVID-19 attributable deaths is also consistent with models that predict substantial age-shifts in transmission dynamics. However, in this case age-shifts in mobility levels are also expected, and we found no evidence for such changes in two independent mobility trend data sets. We further compared model outputs to the number of daily reported COVID-19 cases in each state and metropolitan area, and find that the ratio of estimated, actual cases to reported cases decreases substantially over time (Supplementary Text S7). This suggests that increased testing and increased awareness and test-seeking of individuals aged 20-49 could explain the observed shifts in the age composition of reported cases over the past months [4, 5, 3, 27], because infections among younger individuals are more frequently associated with no or mild symptoms than in older individuals [17, 28]. Fourth, the COVID-19 epidemic is more granular than considered in our spatial modelling approach. Substantial heterogeneity in disease transmission exists at county level [29], and our situation analyses by state and metropolitan areas need to be interpreted as averages. Fifth, the contact and infection model also falls short to account for population structure other than age, such as household settings, where attack rates have been estimated to be substantially higher than in non-household settings [30]. It is possible that we over-estimated the impact of re-opening kindergartens and elementary schools on transmission dynamics. In line with this possibility, contact tracing in elementary schools and further data from countries that have re-opened schools have provided no evidence for substantial transmission in schools, nor increased community-level infection rates [23, 31], although most reports stem from locations with no resurgent epidemics.

## 3 Conclusions

This study provides evidence that the resurgent COVID-19 epidemics in the United States are driven by adults aged 20-49. By August 17, 2020, an estimated 62.7% [60.1%-65.1%] of SARS-CoV-2 infections originated from adults aged 20-49 whereas less than 2% originated from children aged 0-9. We find heterogeneity in age-specific reproduction numbers across locations, with highest reproduction numbers from individuals aged 35-49, followed by individuals aged 20-34. We find no evidence for substantial shifts in contact and transmission dynamics between age groups over time. This suggests that working adults who need to support themselves and their families have been driving the resurging epidemics in the United States. Re-opening kindergartens and elementary schools is essential, but are predicted to facilitate the spread of SARS-Cov-2 in areas with sustained community-wide transmission from adults. This study indicates that targeting interventions at adults aged 20-49 could bring resurgent epidemics under control, avert deaths, and facilitate the safe re-opening of schools.

## 4 Data

The national-level, aggregate mobility data used in this study are described in Supplementary Text S1. The agespecific COVID-19 attributable mortality data used in this study are described in Supplementary Text S2.

## 5 Methods

The contact-and-infection model and further methods are described in Supplementary Text S3.

## 6 Location-specific COVID-19 situation reports

Detailed situation reports for the 37 states and metropolitan areas evaluated in this study are in Supplementary Text S7.

## 7 Comparison to external contact data and COVID-19 seroprevalence data

To gain further insights into the model outputs, we reviewed data from contact surveys during the pandemic, and from several large-scale COVID-19 seroprevalence surveys in the United States. The model outputs are compared to the data from contact surveys in Supplementary Text S4, and to the COVID-19 seroprevalence survey data in Supplementary Text S5.

## 8 Sensitivity analyses

Sensitivity analyses are presented in Supplementary Text S6.

## Data Availability

The COVID-19 mortality data used in this study are available on GitHub, https://github.com/ImperialCollegeLondon/US-covid19-agespecific-mortality-data, under the Creative Commons Attribution 4.0 International Public License. Code and further data are available on Github, https://github.com/ImperialCollegeLondon/covid19model, under the MIT License. 

https://github.com/ImperialCollegeLondon/US-covid19-agespecific-mortality-data

https://github.com/ImperialCollegeLondon/covid19model

## 9 Acknowledgements

This study was supported by the Imperial College COVID-19 Response Fund, the Imperial College Research Computing Service DOI:10.14469/hpc/2232, the Bill & Melinda Gates Foundation, and the EPSRC through the EPSRC Centre for Doctoral Training in Modern Statistics and Statistical Machine Learning at Imperial and Oxford, the UK Medical Research Council under a concordat with the UK Department for International Development, the NIHR Health Protection Research Unit in Modelling Methodology and Community Jameel. We would like to thank Microsoft and Amazon for providing cloud computing services.

## Contributors

OR conceived the study. AG, SM, SF, SB, NF, OR oversaw the study. MM, DH, SBe, ST, YC, McM, MH, HZ, ABe, OR oversaw and performed data collection. MM, ABl, XX, OR lead the analysis. VCB, HC, SF, JIH, TM, AG, HJTU, MV, SW, SM contributed to the analysis. All authors discussed the results and contributed to the revision of the final manuscript.

## Contributors, Imperial College COVID-19 Response Team

We would like to thank the Imperial College COVID-19 Response Team for their insightful comments, Kylie E C Ainslie, Marc Baguelin, Adhiratha Boonyasiri, Olivia Boyd, Lorenzo Cattarino, Laura V Cooper, Zulma Cucunubá, Gina Cuomo-Dannenburg, Bimandra Djaafara, Ilaria Dorigatti, Sabine L van Elsland, Richard FitzJohn, Katy A M Gaythorpe, Lily Geidelberg, William D. Green, Arran Hamlet, Wes Hinsley, Ben Jeffrey, Edward Knock, Daniel Laydon, Gemma Nedjati-Gilani, Pierre Nouvellet, Kris V Parag, Igor Siveroni, Hayley A Thompson, Robert Verity, Caroline E. Walters, Haowei Wang, Yuanrong Wang, Oliver J Watson, Peter Winskill, Charles Whittaker, Patrick GT Walker, Christl A. Donnelly, Lucy Okell, Sangeeta Bhatia, Nicholas F. Brazeau, Oliver D Eales, David Haw, Natsuko Imai, Elita Jauneikaite, John Lees, Andria Mousa, Daniela Olivera, Janetta Skarp, Lilith Whittles

## Supplementary Tables and Figures

**Table S1:** Posterior probability that the weekly age-specific reproduction number is smaller than 1, as of the week starting on August 17, 2020. Posterior mean estimates for each location (state or metropolitan area) and age group in percent. Epidemiologic classifications are: all age-specific reproduction numbers consistently < 1 in last 4 weeks (*•*), reproduction numbers > 1 after a minimum 4-week period of reproduction numbers < 1 for all age groups (*•*), reproduction numbers > 1 from ages 35-49 in last 4 weeks, and no period minimum 4-week period of reproduction numbers < 1 (*•*), reproduction numbers > 1 from ages 35-49 and other age groups in last 4 weeks, and no period minimum 4-week period of reproduction numbers < 1 (*•*).

| Location | Age of infectious individuals (years) |  |  |  |  |  |  |  | Epidemiological classification |
| --- | --- | --- | --- | --- | --- | --- | --- | --- | --- |
|  | Overall | [0 – 9] | [10 – 19] | [20 – 34] | [35 – 49] | [50 – 64] | [65 – 79] | 80+ |  |
| All locations | 99.7 | 100.0 | 94.2 | 76.4 | 0.1 | 100.0 | 100.0 | 100.0 | ● |
| Alabama | 99.9 | 100.0 | 100.0 | 94.8 | 0.0 | 100.0 | 100.0 | 100.0 | ● |
| Alaska | - | - | - | - | - | - | - | - | - |
| Arizona | 100.0 | 100.0 | 99.8 | 99.9 | 80.7 | 100.0 | 100.0 | 100.0 | ● |
| Arkansas | - | - | - | - | - | - | - | - | - |
| California | 99.7 | 100.0 | 86.9 | 26.7 | 0.0 | 100.0 | 100.0 | 100.0 | ● |
| Colorado | 100.0 | 100.0 | 100.0 | 100.0 | 1.1 | 100.0 | 100.0 | 100.0 | ● |
| Connecticut | 100.0 | 100.0 | 100.0 | 100.0 | 100.0 | 100.0 | 100.0 | 100.0 | ● |
| Delaware | 100.0 | 100.0 | 100.0 | 100.0 | 25.2 | 100.0 | 100.0 | 100.0 | ● |
| District of Columbia | 100.0 | 100.0 | 100.0 | 99.9 | 99.8 | 100.0 | 100.0 | 100.0 | ● |
| Florida | 100.0 | 100.0 | 94.2 | 81.3 | 16.8 | 100.0 | 100.0 | 100.0 | ● |
| Georgia | 99.9 | 100.0 | 85.8 | 85.7 | 2.0 | 100.0 | 100.0 | 100.0 | ● |
| Hawaii | - | - | - | - | - | - | - | - | - |
| Idaho | 15.1 | 100.0 | 66.0 | 4.5 | 0.0 | 18.9 | 100.0 | 100.0 | ● |
| Illinois | 100.0 | 100.0 | 100.0 | 100.0 | 9.9 | 100.0 | 100.0 | 100.0 | ● |
| Indiana | 100.0 | 100.0 | 100.0 | 99.4 | 0.0 | 100.0 | 100.0 | 100.0 | ● |
| Iowa | 99.8 | 99.0 | 100.0 | 99.6 | 0.0 | 99.7 | 100.0 | 100.0 | ● |
| Kansas | - | - | - | - | - | - | - | - | - |
| Kentucky | 92.1 | 100.0 | 99.6 | 13.2 | 0.0 | 99.7 | 100.0 | 100.0 | ● |
| Louisiana | 99.5 | 100.0 | 90.2 | 94.7 | 26.2 | 99.9 | 100.0 | 100.0 | ● |
| Maine | - | - | - | - | - | - | - | - | - |
| Maryland | 100.0 | 100.0 | 100.0 | 100.0 | 1.0 | 100.0 | 100.0 | 100.0 | ● |
| Massachusetts | 100.0 | 100.0 | 100.0 | 100.0 | 32.4 | 100.0 | 100.0 | 100.0 | ● |
| Michigan | 100.0 | 100.0 | 100.0 | 98.8 | 3.6 | 100.0 | 100.0 | 100.0 | ● |
| Minnesota | - | - | - | - | - | - | - | - | - |
| Mississippi | 100.0 | 100.0 | 100.0 | 99.5 | 25.4 | 100.0 | 100.0 | 100.0 | ● |
| Missouri | 98.4 | 99.8 | 99.9 | 88.1 | 0.0 | 98.3 | 100.0 | 100.0 | ● |
| Montana | - | - | - | - | - | - | - | - | - |
| Nebraska | - | - | - | - | - | - | - | - | - |
| Nevada | 97.3 | 99.9 | 4.3 | 34.0 | 11.3 | 100.0 | 100.0 | 100.0 | ● |
| New Hampshire | 100.0 | 92.5 | 100.0 | 100.0 | 41.9 | 100.0 | 100.0 | 100.0 | ● |
| New Jersey | 100.0 | 100.0 | 100.0 | 100.0 | 98.9 | 100.0 | 100.0 | 100.0 | ● |
| New Mexico | 100.0 | 94.9 | 55.1 | 92.5 | 0.0 | 100.0 | 100.0 | 100.0 | ● |
| New York | - | - | - | - | - | - | - | - | - |
| New York City | 100.0 | 100.0 | 100.0 | 100.0 | 89.6 | 100.0 | 100.0 | 100.0 | ● |
| North Carolina | 91.9 | 99.9 | 100.0 | 40.1 | 0.0 | 97.1 | 100.0 | 100.0 | ● |
| North Dakota | - | - | - | - | - | - | - | - | - |
| Ohio | - | - | - | - | - | - | - | - | - |
| Oklahoma | 69.5 | 100.0 | 88.7 | 24.4 | 0.0 | 98.0 | 100.0 | 100.0 | ● |
| Oregon | 5.8 | 100.0 | 24.1 | 0.2 | 0.0 | 44.2 | 100.0 | 100.0 | ● |
| Pennsylvania | 99.8 | 98.9 | 100.0 | 96.7 | 0.0 | 99.7 | 100.0 | 100.0 | ● |
| Rhode Island | 100.0 | 100.0 | 100.0 | 100.0 | 41.1 | 100.0 | 100.0 | 100.0 | ● |
| South Carolina | 99.5 | 100.0 | 97.5 | 36.7 | 0.0 | 100.0 | 100.0 | 100.0 | ● |
| South Dakota | - | - | - | - | - | - | - | - | - |
| Tennessee | 54.9 | 100.0 | 78.9 | 1.0 | 0.0 | 99.0 | 100.0 | 100.0 | ● |
| Texas | 95.9 | 100.0 | 66.0 | 66.9 | 1.7 | 99.9 | 100.0 | 100.0 | ● |
| Utah | 21.9 | 98.5 | 81.0 | 19.5 | 0.0 | 99.9 | 100.0 | 100.0 | ● |
| Vermont | - | - | - | - | - | - | - | - | - |
| Virginia | 100.0 | 100.0 | 100.0 | 99.5 | 0.0 | 100.0 | 100.0 | 100.0 | ● |
| Washington | 67.8 | 100.0 | 7.7 | 7.6 | 0.1 | 95.2 | 100.0 | 100.0 | ● |
| West Virginia | - | - | - | - | - | - | - | - | - |
| Wisconsin | 96.4 | 100.0 | 100.0 | 58.9 | 0.0 | 95.2 | 100.0 | 100.0 | ● |
| Wyoming | - | - | - | - | - | - | - | - | - |

**Table S2:** Estimated age-specific weekly effective reproduction number, as of the week starting on August 17, 2020. Posterior median estimates for each location (state or metropolitan area) and age group, along with 95% credible intervals.

| Location | Age of infectious individuals (years) |  |  |  |  |  |  |  |
| --- | --- | --- | --- | --- | --- | --- | --- | --- |
|  | Overall | [0 – 9] | [10 – 19] | [20 – 34] | [35 – 49] | [50 – 64] | [65 – 79] | 80+ |
| All locations | 0.91 [0.86-0.97] | 0.58 [0.52-0.64] | 0.93 [0.86-1.02] | 0.98 [0.92-1.05] | 1.10 [1.04-1.17] | 0.85 [0.80-0.91] | 0.50 [0.45-0.57] | 0.26 [0.24-0.30] |
| Alabama | 0.90 [0.84-0.96] | 0.56 [0.43-0.72] | 0.68 [0.59-0.81] | 0.94 [0.86-1.01] | 1.16 [1.09-1.23] | 0.87 [0.81-0.93] | 0.48 [0.42-0.54] | 0.25 [0.22-0.28] |
| Alaska | - | - | - | - | - | - | - | - |
| Arizona | 0.81 [0.73-0.87] | 0.51 [0.38-0.65] | 0.88 [0.79-0.97] | 0.90 [0.81-0.97] | 0.96 [0.88-1.04] | 0.74 [0.68-0.80] | 0.44 [0.39-0.50] | 0.23 [0.21-0.26] |
| Arkansas | - | - | - | - | - | - | - | - |
| California | 0.96 [0.93-0.99] | 0.60 [0.49-0.73] | 0.97 [0.92-1.02] | 1.01 [0.97-1.05] | 1.15 [1.11-1.19] | 0.91 [0.87-0.95] | 0.50 [0.45-0.56] | 0.26 [0.24-0.29] |
| Colorado | 0.86 [0.83-0.89] | 0.65 [0.52-0.80] | 0.86 [0.82-0.91] | 0.89 [0.85-0.92] | 1.04 [1.01-1.09] | 0.80 [0.76-0.85] | 0.55 [0.49-0.62] | 0.28 [0.25-0.32] |
| Connecticut | 0.71 [0.68-0.75] | 0.55 [0.41-0.70] | 0.55 [0.52-0.60] | 0.71 [0.68-0.76] | 0.89 [0.86-0.94] | 0.76 [0.73-0.81] | 0.51 [0.46-0.56] | 0.27 [0.25-0.30] |
| Delaware | 0.82 [0.78-0.86] | 0.53 [0.39-0.69] | 0.74 [0.70-0.82] | 0.91 [0.87-0.96] | 1.02 [0.97-1.06] | 0.80 [0.76-0.84] | 0.46 [0.41-0.52] | 0.24 [0.22-0.27] |
| District of Columbia | 0.78 [0.74-0.84] | 0.45 [0.32-0.61] | 0.70 [0.66-0.77] | 0.89 [0.84-0.96] | 0.92 [0.87-0.97] | 0.64 [0.60-0.70] | 0.42 [0.38-0.49] | 0.22 [0.19-0.25] |
| Florida | 0.86 [0.78-0.94] | 0.49 [0.39-0.59] | 0.91 [0.81-1.02] | 0.96 [0.87-1.05] | 1.04 [0.95-1.14] | 0.79 [0.72-0.86] | 0.47 [0.42-0.54] | 0.25 [0.22-0.28] |
| Georgia | 0.90 [0.84-0.96] | 0.43 [0.35-0.51] | 0.94 [0.85-1.05] | 0.96 [0.89-1.03] | 1.07 [1.01-1.14] | 0.84 [0.78-0.89] | 0.52 [0.47-0.58] | 0.27 [0.25-0.30] |
| Hawaii | - | - | - | - | - | - | - | - |
| Idaho | 1.08 [0.93-1.23] | 0.49 [0.36-0.67] | 0.96 [0.79-1.17] | 1.13 [0.98-1.30] | 1.32 [1.14-1.51] | 1.07 [0.91-1.25] | 0.68 [0.55-0.84] | 0.35 [0.29-0.43] |
| Illinois | 0.83 [0.81-0.86] | 0.71 [0.58-0.86] | 0.83 [0.80-0.86] | 0.86 [0.83-0.89] | 1.02 [0.99-1.06] | 0.80 [0.77-0.84] | 0.48 [0.44-0.53] | 0.26 [0.23-0.28] |
| Indiana | 0.90 [0.86-0.95] | 0.65 [0.52-0.81] | 0.75 [0.70-0.82] | 0.93 [0.88-0.98] | 1.12 [1.07-1.19] | 0.88 [0.83-0.94] | 0.54 [0.48-0.61] | 0.28 [0.25-0.32] |
| Iowa | 0.93 [0.90-0.97] | 0.72 [0.52-0.95] | 0.66 [0.62-0.75] | 0.92 [0.86-0.98] | 1.20 [1.15-1.26] | 0.92 [0.88-0.97] | 0.56 [0.50-0.63] | 0.29 [0.26-0.33] |
| Kansas | - | - | - | - | - | - | - | - |
| Kentucky | 0.97 [0.92-1.01] | 0.60 [0.44-0.79] | 0.82 [0.74-0.95] | 1.03 [0.97-1.09] | 1.20 [1.14-1.26] | 0.92 [0.87-0.98] | 0.51 [0.45-0.58] | 0.27 [0.24-0.30] |
| Louisiana | 0.84 [0.74-0.96] | 0.47 [0.37-0.58] | 0.88 [0.73-1.06] | 0.90 [0.78-1.03] | 1.04 [0.92-1.16] | 0.82 [0.72-0.92] | 0.50 [0.43-0.58] | 0.26 [0.23-0.30] |
| Maine | - | - | - | - | - | - | - | - |
| Maryland | 0.83 [0.80-0.86] | 0.65 [0.51-0.82] | 0.68 [0.65-0.72] | 0.86 [0.82-0.89] | 1.04 [1.01-1.08] | 0.83 [0.80-0.86] | 0.46 [0.41-0.50] | 0.24 [0.22-0.27] |
| Massachusetts | 0.81 [0.75-0.85] | 0.60 [0.44-0.77] | 0.63 [0.59-0.69] | 0.80 [0.75-0.86] | 1.01 [0.95-1.07] | 0.84 [0.79-0.90] | 0.60 [0.55-0.66] | 0.32 [0.29-0.35] |
| Michigan | 0.86 [0.82-0.91] | 0.63 [0.51-0.76] | 0.78 [0.73-0.84] | 0.94 [0.89-0.99] | 1.05 [1.00-1.11] | 0.84 [0.79-0.89] | 0.51 [0.45-0.58] | 0.27 [0.24-0.30] |
| Minnesota | - | - | - | - | - | - | - | - |
| Mississippi | 0.82 [0.74-0.89] | 0.49 [0.39-0.62] | 0.76 [0.66-0.87] | 0.88 [0.80-0.97] | 1.03 [0.94-1.12] | 0.77 [0.70-0.84] | 0.41 [0.36-0.47] | 0.22 [0.19-0.25] |
| Missouri | 0.95 [0.92-0.99] | 0.67 [0.52-0.87] | 0.83 [0.77-0.92] | 0.97 [0.91-1.02] | 1.21 [1.15-1.26] | 0.94 [0.89-1.00] | 0.55 [0.49-0.62] | 0.29 [0.26-0.33] |
| Montana | - | - | - | - | - | - | - | - |
| Nebraska | - | - | - | - | - | - | - | - |
| Nevada | 0.92 [0.84-1.00] | 0.68 [0.51-0.88] | 1.09 [0.99-1.21] | 1.02 [0.93-1.11] | 1.05 [0.97-1.14] | 0.80 [0.73-0.87] | 0.53 [0.47-0.60] | 0.27 [0.24-0.31] |
| New Hampshire | 0.81 [0.73-0.87] | 0.78 [0.53-1.09] | 0.67 [0.61-0.76] | 0.81 [0.72-0.89] | 1.01 [0.91-1.09] | 0.80 [0.72-0.87] | 0.51 [0.43-0.59] | 0.26 [0.23-0.30] |
| New Jersey | 0.77 [0.72-0.81] | 0.49 [0.38-0.61] | 0.75 [0.71-0.80] | 0.84 [0.80-0.89] | 0.94 [0.89-0.99] | 0.75 [0.71-0.79] | 0.46 [0.42-0.51] | 0.24 [0.22-0.27] |
| New Mexico | 0.90 [0.87-0.93] | 0.80 [0.60-1.04] | 1.00 [0.96-1.04] | 0.97 [0.94-1.01] | 1.08 [1.04-1.11] | 0.80 [0.76-0.84] | 0.47 [0.42-0.53] | 0.24 [0.22-0.28] |
| New York | - | - | - | - | - | - | - | - |
| New York City | 0.77 [0.70-0.86] | 0.36 [0.28-0.45] | 0.73 [0.64-0.85] | 0.83 [0.74-0.93] | 0.94 [0.85-1.03] | 0.76 [0.69-0.84] | 0.49 [0.43-0.55] | 0.26 [0.23-0.29] |
| North Carolina | 0.98 [0.94-1.01] | 0.68 [0.52-0.87] | 0.76 [0.72-0.86] | 1.01 [0.96-1.05] | 1.23 [1.19-1.28] | 0.96 [0.91-1.00] | 0.60 [0.54-0.67] | 0.31 [0.28-0.35] |
| North Dakota | - | - | - | - | - | - | - | - |
| Ohio | - | - | - | - | - | - | - | - |
| Oklahoma | 0.98 [0.88-1.08] | 0.54 [0.41-0.71] | 0.89 [0.74-1.07] | 1.04 [0.93-1.16] | 1.21 [1.09-1.33] | 0.90 [0.81-0.99] | 0.52 [0.44-0.61] | 0.27 [0.23-0.31] |
| Oregon | 1.06 [0.98-1.14] | 0.55 [0.40-0.72] | 1.04 [0.94-1.15] | 1.12 [1.04-1.20] | 1.28 [1.19-1.38] | 1.01 [0.92-1.11] | 0.62 [0.54-0.72] | 0.31 [0.27-0.36] |
| Pennsylvania | 0.90 [0.85-0.96] | 0.79 [0.64-0.97] | 0.73 [0.68-0.78] | 0.94 [0.88-1.00] | 1.14 [1.06-1.21] | 0.90 [0.83-0.96] | 0.53 [0.47-0.61] | 0.28 [0.25-0.32] |
| Rhode Island | 0.79 [0.75-0.82] | 0.63 [0.47-0.83] | 0.53 [0.50-0.58] | 0.76 [0.71-0.80] | 1.01 [0.96-1.05] | 0.84 [0.80-0.88] | 0.56 [0.51-0.61] | 0.29 [0.27-0.32] |
| South Carolina | 0.92 [0.86-0.98] | 0.46 [0.36-0.57] | 0.88 [0.78-1.00] | 1.01 [0.94-1.08] | 1.13 [1.06-1.21] | 0.85 [0.79-0.91] | 0.48 [0.42-0.55] | 0.25 [0.22-0.28] |
| South Dakota | - | - | - | - | - | - | - | - |
| Tennessee | 1.00 [0.95-1.04] | 0.49 [0.36-0.65] | 0.95 [0.84-1.07] | 1.07 [1.01-1.13] | 1.22 [1.16-1.29] | 0.93 [0.88-0.99] | 0.56 [0.50-0.64] | 0.29 [0.26-0.33] |
| Texas | 0.93 [0.85-1.01] | 0.60 [0.48-0.74] | 0.98 [0.87-1.10] | 0.98 [0.89-1.07] | 1.10 [1.01-1.20] | 0.87 [0.79-0.95] | 0.50 [0.44-0.57] | 0.26 [0.23-0.30] |
| Utah | 1.01 [0.98-1.04] | 0.71 [0.50-0.97] | 0.95 [0.87-1.07] | 1.02 [0.96-1.07] | 1.25 [1.20-1.31] | 0.91 [0.87-0.96] | 0.43 [0.38-0.49] | 0.23 [0.20-0.25] |
| Vermont | - | - | - | - | - | - | - | - |
| Virginia | 0.93 [0.90-0.95] | 0.66 [0.53-0.83] | 0.68 [0.65-0.75] | 0.95 [0.92-0.99] | 1.16 [1.13-1.20] | 0.92 [0.89-0.96] | 0.51 [0.46-0.57] | 0.27 [0.24-0.30] |
| Washington | 0.98 [0.90-1.06] | 0.33 [0.29-0.37] | 1.07 [0.97-1.16] | 1.06 [0.98-1.14] | 1.14 [1.04-1.24] | 0.92 [0.84-1.02] | 0.64 [0.55-0.75] | 0.33 [0.28-0.38] |
| West Virginia | - | - | - | - | - | - | - | - |
| Wisconsin | 0.96 [0.92-1.00] | 0.68 [0.51-0.87] | 0.83 [0.78-0.91] | 0.99 [0.94-1.05] | 1.18 [1.13-1.25] | 0.95 [0.90-1.01] | 0.61 [0.54-0.69] | 0.32 [0.28-0.36] |
| Wyoming | - | - | - | - | - | - | - | - |

**Table S3:** Cumulative age-specific attack rates, as of August 23, 2020. Posterior median estimates are shown in percent, along with 95% credible intervals.

| Location | Age of infectious individuals (years) |  |  |  |  |  |  |  |
| --- | --- | --- | --- | --- | --- | --- | --- | --- |
|  | Overall | [0 – 9] | [10 – 19] | [20 – 34] | [35 – 49] | [50 – 64] | [65 – 79] | 80+ |
| Alabama | 13.68 [9.61-19.10] | 2.12 [1.17-3.83] | 6.57 [4.49-9.49] | 17.42 [12.27-24.14] | 23.35 [16.39-32.15] | 15.64 [10.82-21.98] | 10.84 [6.61-17.99] | 7.35 [4.49-12.20] |
| Alaska | - | - | - | - | - | - | - | - |
| Arizona | 24.61 [19.11-31.41] | 3.73 [2.18-6.39] | 20.32 [15.82-25.37] | 34.30 [26.61-43.19] | 36.87 [28.64-46.30] | 25.99 [19.84-33.37] | 18.00 [12.01-28.03] | 11.95 [7.93-18.87] |
| Arkansas | - | - | - | - | - | - | - | - |
| California | 9.21 [6.24-13.01] | 1.28 [0.70-2.24] | 6.69 [4.57-9.46] | 11.76 [8.01-16.60] | 13.67 [9.32-19.23] | 10.04 [6.78-14.31] | 7.62 [4.47-12.67] | 4.77 [2.81-7.93] |
| Colorado | 9.82 [6.37-14.10] | 1.69 [0.91-2.95] | 7.99 [5.23-11.31] | 12.11 [7.89-17.26] | 14.06 [9.12-20.03] | 9.90 [6.36-14.22] | 9.02 [5.11-15.53] | 6.27 [3.57-10.75] |
| Connecticut | 28.67 [17.54-39.65] | 5.83 [3.18-9.75] | 16.17 [9.79-23.25] | 33.75 [20.81-46.56] | 41.97 [26.44-56.26] | 33.12 [20.19-45.79] | 28.75 [15.95-46.88] | 17.68 [9.55-29.99] |
| Delaware | 17.34 [10.64-25.26] | 2.88 [1.44-5.42] | 11.03 [6.77-16.14] | 23.83 [14.80-34.01] | 27.20 [16.84-38.62] | 18.79 [11.41-27.54] | 12.99 [7.02-22.40] | 9.17 [4.92-15.91] |
| District of Columbia | 26.89 [19.96-35.46] | 5.38 [3.12-9.15] | 17.09 [12.43-23.18] | 34.52 [25.68-44.97] | 35.92 [26.99-46.48] | 26.05 [19.12-34.76] | 22.50 [14.26-35.41] | 14.74 [9.24-23.70] |
| Florida | 20.16 [14.32-27.30] | 2.95 [1.72-5.01] | 18.19 [13.03-24.31] | 29.06 [20.74-38.90] | 30.36 [21.70-40.31] | 20.26 [14.28-27.57] | 13.92 [8.62-22.51] | 7.92 [4.89-12.96] |
| Georgia | 14.68 [10.16-19.74] | 1.50 [0.86-2.49] | 11.10 [7.69-15.07] | 19.25 [13.42-25.75] | 21.18 [14.89-28.17] | 15.46 [10.73-20.85] | 13.33 [8.02-21.41] | 9.94 [5.96-16.01] |
| Hawaii | - | - | - | - | - | - | - | - |
| Idaho | 5.34 [2.72-8.90] | 0.53 [0.23-1.11] | 3.14 [1.63-5.27] | 7.05 [3.67-11.63] | 8.42 [4.29-13.98] | 6.19 [3.11-10.52] | 4.91 [2.24-9.71] | 3.41 [1.56-6.68] |
| Illinois | 17.02 [12.47-23.58] | 3.54 [2.07-6.13] | 12.66 [9.19-17.46] | 21.37 [15.63-29.32] | 25.57 [18.74-34.82] | 18.18 [13.21-25.21] | 14.56 [9.20-23.91] | 9.21 [5.84-15.26] |
| Indiana | 11.43 [6.34-17.02] | 1.84 [0.88-3.48] | 6.50 [3.64-9.97] | 14.40 [8.08-21.54] | 18.17 [10.14-26.91] | 12.86 [7.07-19.40] | 10.20 [5.08-17.71] | 6.58 [3.25-11.48] |
| Iowa | 7.61 [4.61-11.19] | 1.30 [0.66-2.55] | 3.32 [2.01-4.98] | 9.22 [5.65-13.46] | 13.30 [8.06-19.44] | 8.91 [5.35-13.15] | 6.98 [3.73-12.11] | 3.94 [2.10-6.87] |
| Kansas | - | - | - | - | - | - | - | - |
| Kentucky | 5.41 [3.20-7.98] | 0.76 [0.37-1.45] | 3.08 [1.83-4.58] | 7.17 [4.28-10.51] | 8.78 [5.19-13.06] | 5.93 [3.48-8.85] | 4.34 [2.25-7.46] | 2.80 [1.45-4.81] |
| Louisiana | 25.41 [17.60-33.69] | 3.79 [2.23-6.31] | 18.94 [13.07-25.25] | 32.71 [22.69-43.06] | 38.04 [26.84-49.00] | 27.87 [19.12-36.89] | 22.15 [13.21-35.15] | 15.57 [9.26-25.15] |
| Maine | - | - | - | - | - | - | - | - |
| Maryland | 16.88 [11.71-23.58] | 3.23 [1.84-5.65] | 9.56 [6.45-13.55] | 20.95 [14.44-28.91] | 26.25 [18.31-35.97] | 19.05 [13.08-26.65] | 14.19 [8.48-23.51] | 9.62 [5.73-16.09] |
| Massachusetts | 27.82 [14.80-39.87] | 5.92 [2.86-10.31] | 15.08 [7.66-22.67] | 30.64 [16.27-43.73] | 40.43 [22.06-55.38] | 31.79 [16.79-45.04] | 30.61 [14.82-51.19] | 19.07 [8.99-33.52] |
| Michigan | 11.27 [7.77-15.64] | 2.02 [1.16-3.53] | 7.85 [5.38-10.99] | 14.97 [10.29-20.68] | 17.27 [11.95-23.79] | 11.93 [8.23-16.56] | 9.24 [5.48-15.43] | 5.80 [3.44-9.66] |
| Minnesota | - | - | - | - | - | - | - | - |
| Mississippi | 22.74 [16.98-29.81] | 3.49 [2.07-5.73] | 13.63 [9.78-18.64] | 30.28 [22.60-39.22] | 36.62 [27.88-46.76] | 25.61 [19.05-33.64] | 17.81 [11.26-28.20] | 12.91 [8.16-20.66] |
| Missouri | 5.90 [3.34-8.93] | 0.96 [0.46-1.84] | 3.59 [2.08-5.49] | 7.40 [4.26-11.10] | 9.66 [5.51-14.55] | 6.56 [3.72-9.99] | 4.86 [2.46-8.64] | 3.02 [1.54-5.37] |
| Montana | - | - | - | - | - | - | - | - |
| Nebraska | - | - | - | - | - | - | - | - |
| Nevada | 15.50 [11.00-21.44] | 2.58 [1.42-4.81] | 14.94 [10.81-20.26] | 21.31 [15.20-29.39] | 21.45 [15.29-29.64] | 14.74 [10.38-20.63] | 12.31 [7.63-20.27] | 8.86 [5.53-14.58] |
| New Hampshire | 4.63 [2.23-9.01] | 1.01 [0.38-2.53] | 2.39 [1.15-4.73] | 5.70 [2.78-11.02] | 7.50 [3.60-14.42] | 4.96 [2.38-9.67] | 4.10 [1.71-9.13] | 2.53 [1.06-5.62] |
| New Jersey | 32.24 [24.37-40.30] | 6.51 [4.04-10.11] | 24.07 [18.16-29.92] | 42.21 [31.99-51.91] | 45.94 [35.23-55.79] | 34.58 [25.84-43.29] | 28.21 [18.11-42.90] | 17.71 [11.21-27.70] |
| New Mexico | 11.22 [8.33-15.28] | 2.32 [1.28-4.13] | 9.24 [6.90-12.42] | 15.57 [11.58-20.97] | 17.65 [13.10-23.93] | 11.27 [8.32-15.37] | 7.69 [4.88-12.39] | 4.98 [3.19-8.05] |
| New York | - | - | - | - | - | - | - | - |
| New York City | 32.26 [23.50-40.74] | 4.64 [2.87-7.33] | 22.93 [16.86-28.82] | 39.86 [29.22-50.40] | 44.84 [33.30-55.40] | 35.78 [26.14-45.11] | 29.96 [18.20-44.31] | 18.87 [11.26-28.86] |
| North Carolina | 7.12 [5.07-9.92] | 1.14 [0.63-2.08] | 3.55 [2.51-5.03] | 8.77 [6.28-12.12] | 11.42 [8.17-15.80] | 8.02 [5.70-11.21] | 6.49 [3.98-10.75] | 4.40 [2.72-7.28] |
| North Dakota | - | - | - | - | - | - | - | - |
| Ohio | - | - | - | - | - | - | - | - |
| Oklahoma | 6.00 [3.83-8.89] | 0.75 [0.39-1.40] | 3.57 [2.27-5.32] | 8.04 [5.19-11.78] | 9.92 [6.35-14.57] | 6.64 [4.23-9.83] | 4.92 [2.76-8.58] | 3.08 [1.73-5.36] |
| Oregon | 3.34 [1.90-5.08] | 0.43 [0.20-0.84] | 2.38 [1.37-3.60] | 4.34 [2.48-6.60] | 5.04 [2.83-7.70] | 3.57 [1.99-5.47] | 2.68 [1.34-4.78] | 1.68 [0.85-2.98] |
| Pennsylvania | 12.36 [6.25-19.97] | 2.82 [1.21-5.55] | 7.05 [3.52-11.67] | 15.46 [7.93-24.93] | 19.81 [10.16-31.46] | 13.56 [6.84-22.03] | 10.45 [4.79-19.74] | 6.06 [2.75-11.49] |
| Rhode Island | 16.76 [10.20-25.25] | 3.43 [1.73-6.22] | 7.28 [4.32-11.49] | 17.97 [10.96-27.12] | 26.27 [16.18-38.37] | 19.88 [11.99-29.78] | 18.20 [9.99-32.07] | 10.54 [5.70-18.96] |
| South Carolina | 13.28 [9.17-18.46] | 1.58 [0.89-2.74] | 9.37 [6.44-13.23] | 18.74 [13.01-25.77] | 21.13 [14.63-29.05] | 13.92 [9.54-19.42] | 9.44 [5.75-15.37] | 6.63 [4.05-10.81] |
| South Dakota | - | - | - | - | - | - | - | - |
| Tennessee | 8.94 [6.62-12.27] | 0.99 [0.56-1.82] | 5.93 [4.27-8.27] | 11.99 [8.84-16.26] | 13.97 [10.34-18.96] | 9.52 [7.00-13.01] | 7.30 [4.65-11.92] | 4.86 [3.13-7.95] |
| Texas | 18.40 [13.76-24.54] | 2.48 [1.46-4.23] | 14.08 [10.49-18.73] | 23.97 [17.94-31.82] | 26.87 [20.21-35.37] | 20.26 [15.04-27.18] | 16.30 [10.49-25.84] | 11.86 [7.63-18.94] |
| Utah | 5.21 [3.50-7.42] | 0.64 [0.32-1.23] | 3.14 [2.10-4.59] | 6.64 [4.49-9.40] | 8.82 [5.85-12.51] | 6.37 [4.21-9.11] | 4.29 [2.57-7.13] | 3.06 [1.83-5.07] |
| Vermont | - | - | - | - | - | - | - | - |
| Virginia | 7.68 [4.64-11.28] | 1.29 [0.66-2.37] | 3.80 [2.34-5.64] | 9.34 [5.72-13.52] | 12.28 [7.47-17.83] | 8.90 [5.35-13.01] | 6.63 [3.57-11.45] | 4.37 [2.36-7.57] |
| Washington | 7.02 [4.32-10.14] | 0.72 [0.39-1.25] | 5.96 [3.72-8.55] | 9.28 [5.76-13.41] | 9.79 [6.07-14.18] | 7.19 [4.41-10.47] | 6.32 [3.35-10.95] | 4.19 [2.24-7.21] |
| West Virginia | - | - | - | - | - | - | - | - |
| Wisconsin | 5.06 [3.31-7.14] | 0.88 [0.47-1.61] | 3.10 [2.04-4.41] | 6.41 [4.21-9.07] | 7.95 [5.19-11.20] | 5.50 [3.56-7.78] | 4.64 [2.69-7.82] | 2.75 [1.60-4.59] |
| Wyoming | - | - | - | - | - | - | - | - |

**Table S4:**
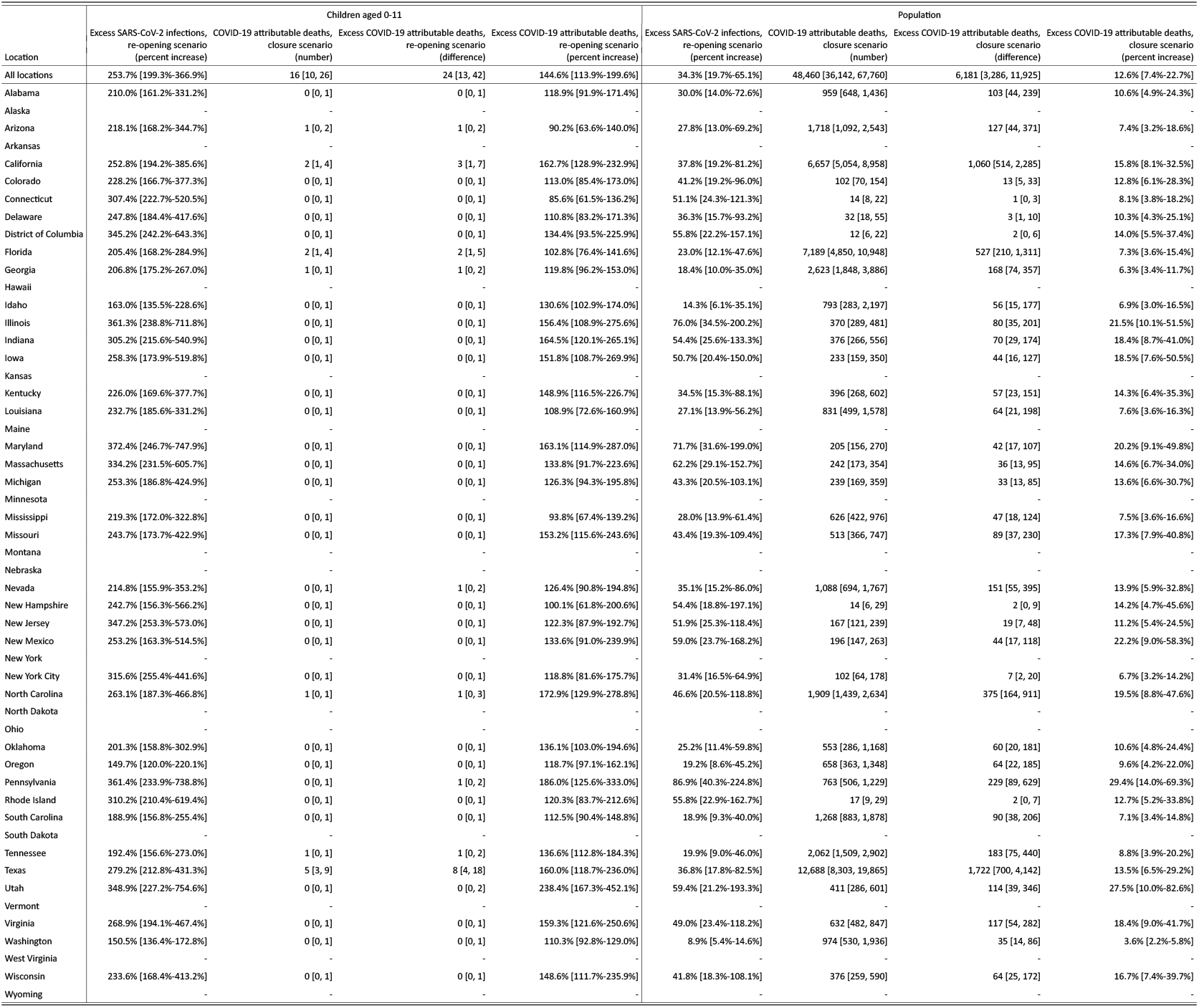
Estimated excess SARS-CoV-2 infections and excess COVID-19 attributable deaths in the central kindergarten and elementary school re-opening scenarios, when compared to continued school closure scenarios. Posterior median estimates for each location (state or metropolitan area), along with 95% credible intervals for the period August 24, 2020 to November 24, 2020. Figures assume a 50% transmission reduction from and to children aged 0-11 due to face mask use and other non-pharmaceutical interventions, see Supplementary Text S3.7.

**Table S5:**
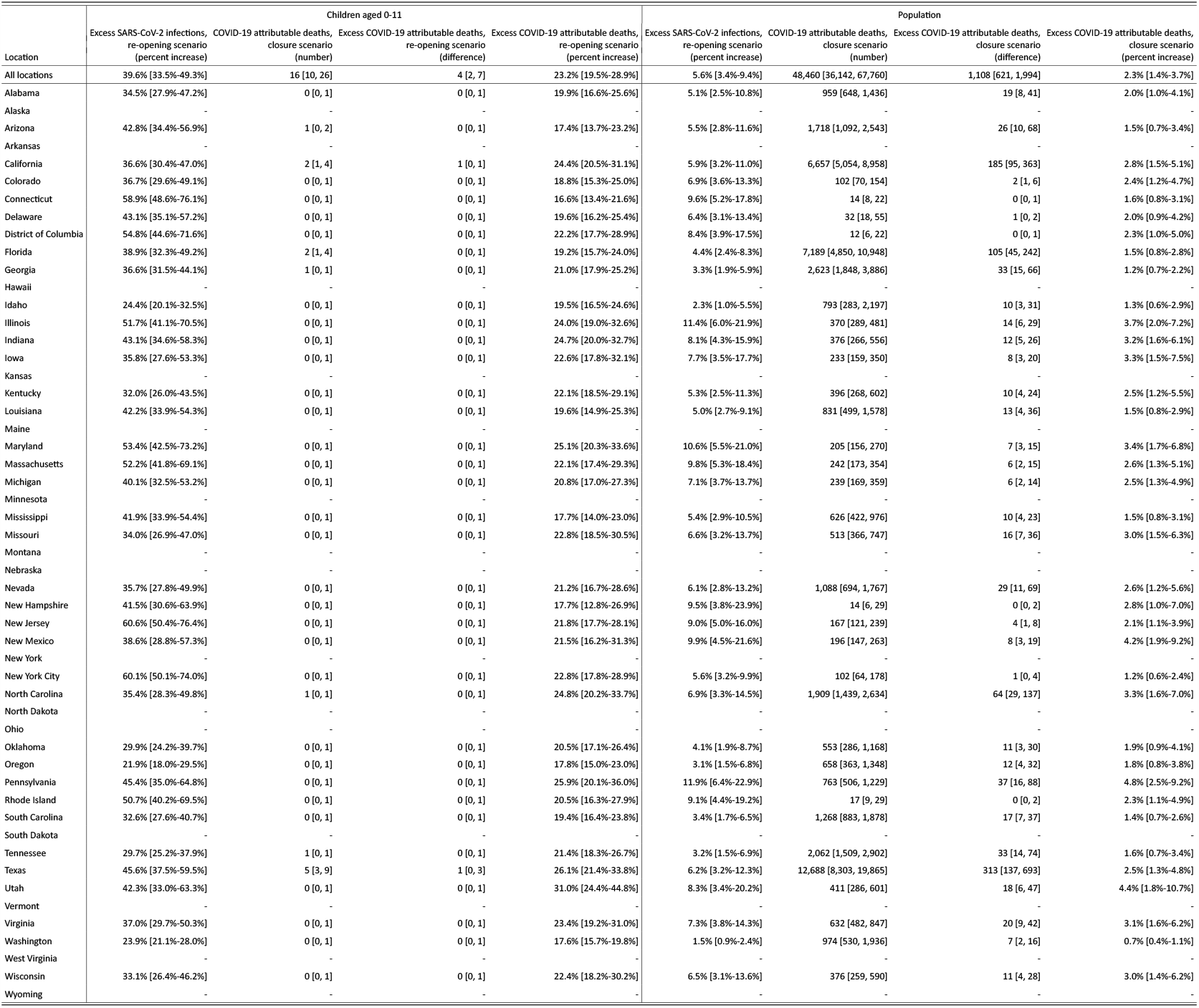
Estimated excess SARS-CoV-2 infections and excess COVID-19 attributable deaths in the kindergarten and elementary school re-opening scenarios with 80% transmission reduction, when compared to continued school closure scenarios. Posterior median estimates for each location (state or metropolitan area), along with 95% credible intervals for the period August 24, 2020 to November 24, 2020. Figures assume a 80% transmission reduction from and to children aged 0-11 due to face mask use and other non-pharmaceutical interventions, see Supplementary Text S3.7.

**Table S6:**
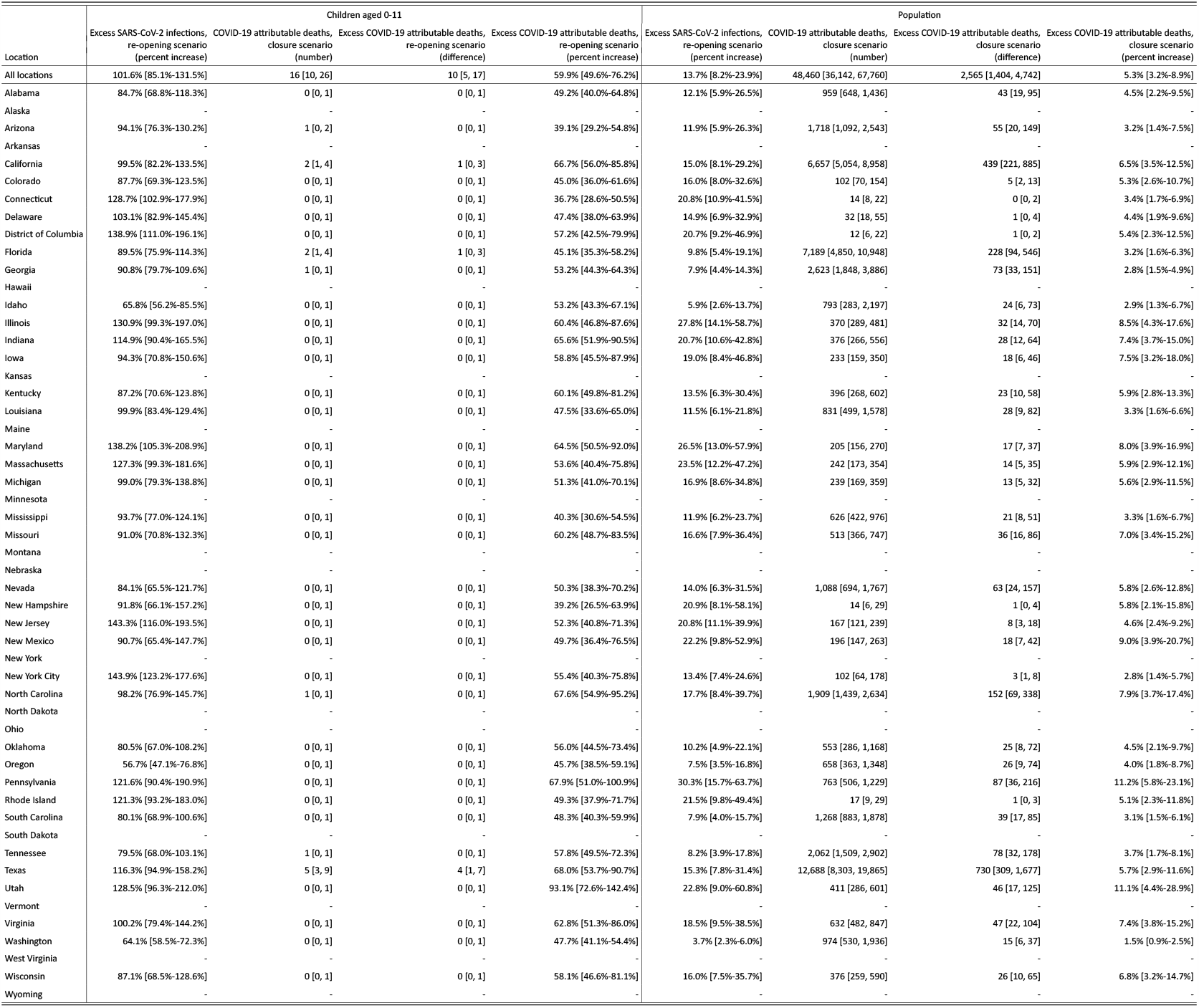
Estimated excess SARS-CoV-2 infections and excess COVID-19 attributable deaths in the kindergarten and elementary school re-opening scenarios with 66% transmission reduction, when compared to continued school closure scenarios. Posterior median estimates for each location (state or metropolitan area), along with 95% credible intervals for the period August 24, 2020 to November 24, 2020. Figures assume a 66% transmission reduction from and to children aged 0-11 due to face mask use and other non-pharmaceutical interventions, see Supplementary Text S3.7.

**Table S7:**
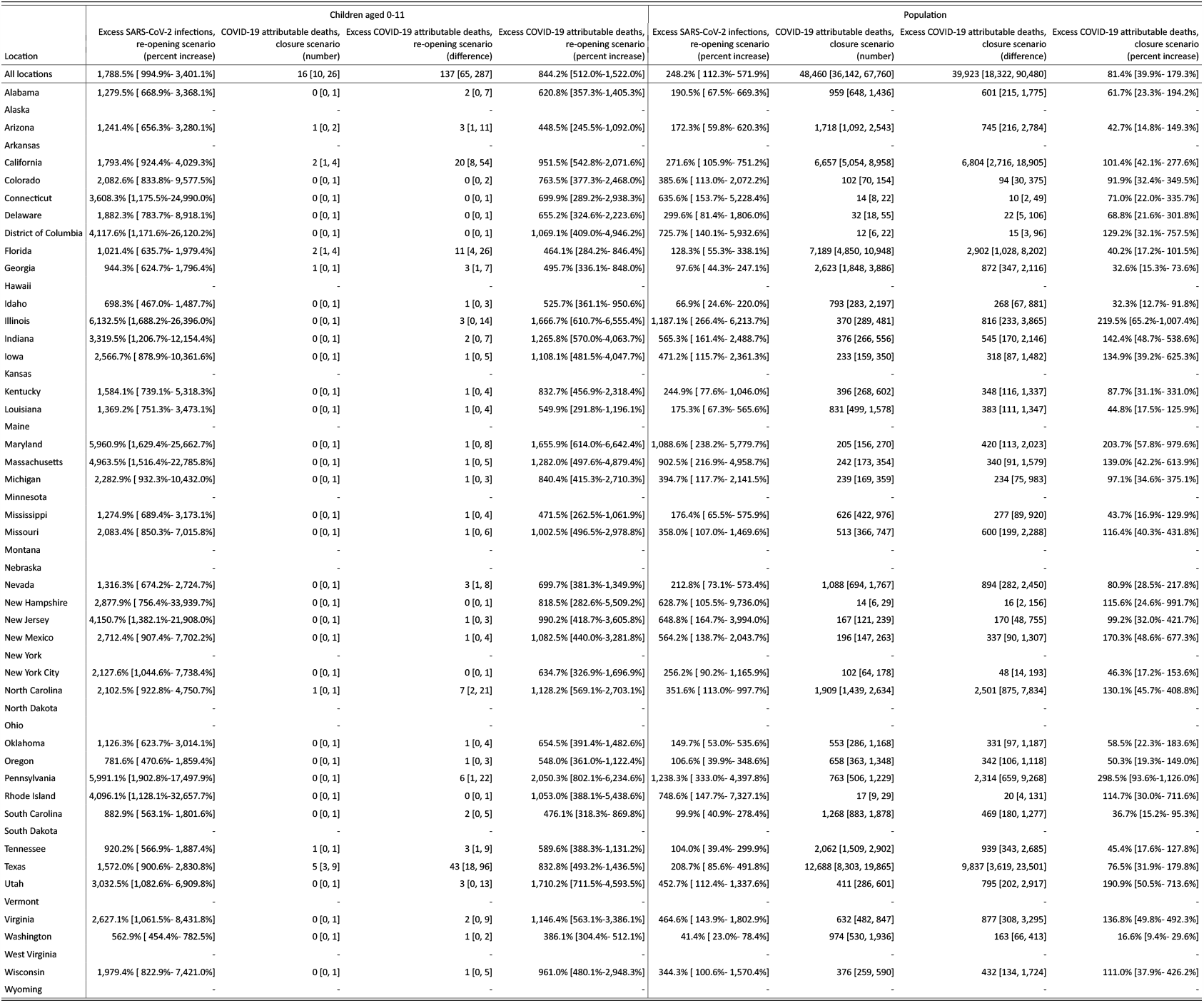
Estimated excess SARS-CoV-2 infections and excess COVID-19 attributable deaths in the kindergarten and elementary school re-opening scenarios with no additional transmission reduction, when compared to continued school closure scenarios. Posterior median estimates for each location (state or metropolitan area), along with 95% credible intervals for the period August 24, 2020 to November 24, 2020. Figures assume no additional transmission reduction from and to children aged 0-11 due to face mask use and other non-pharmaceutical interventions, see Supplementary Text S3.7.

**Figure S1:**
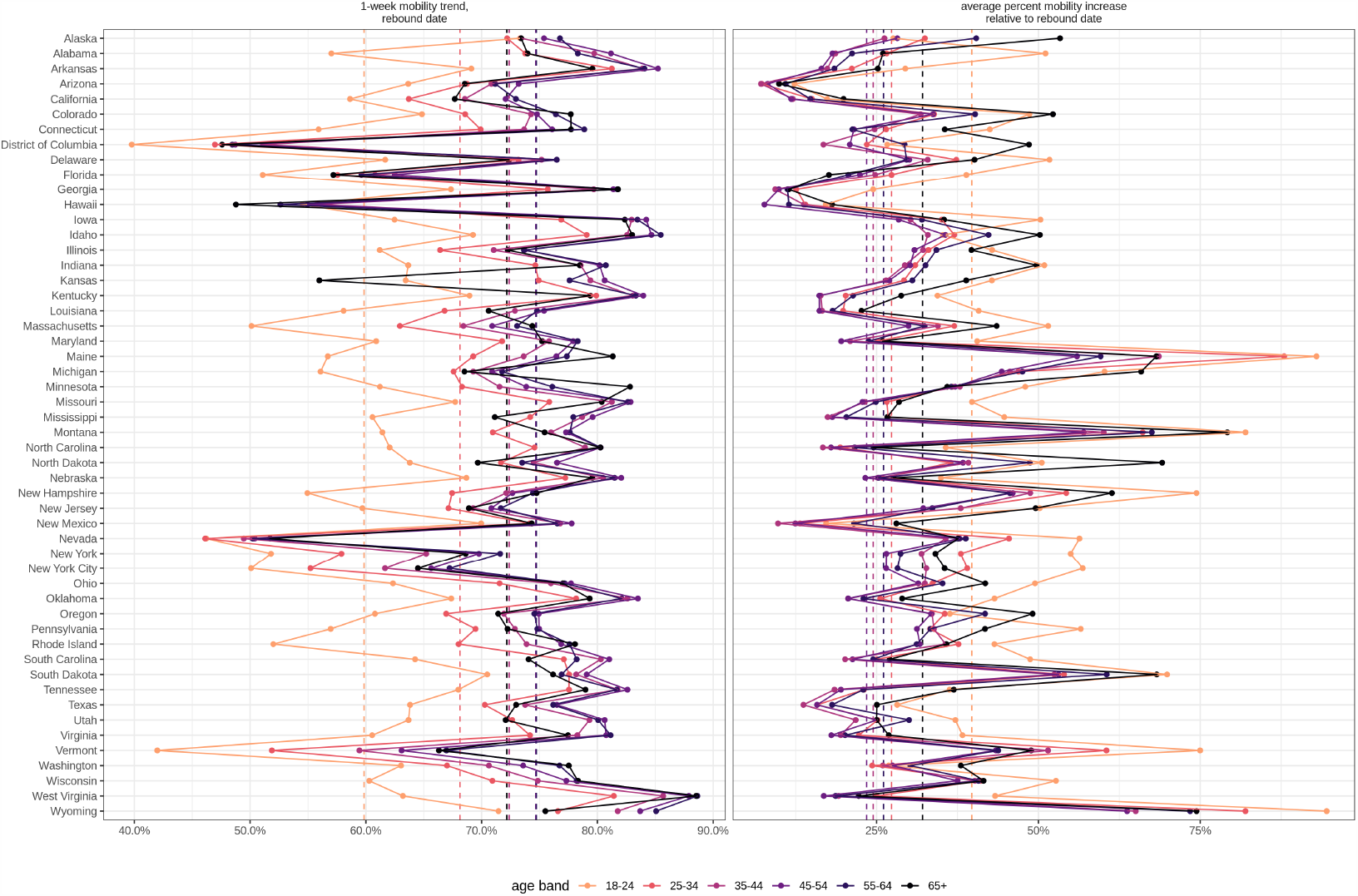
Initial decline and surge in age-specific mobility trends in the United States. (**A**) Longitudinal mobility trends for individuals aged 18 − 24, 25 − 34, 35 − 44, 45 − 54, 55 − 64, 65+ showed an initial decline and a subsequent increase across the United States. Rebound dates were estimated from the time series data, and to have occurred between March 30, 2020 to April 20, 2020. The figure shows age-specific mobility trends relative to the baseline period February 03 to February 09, 2020 for each location (state or metropolitan area). The 1-week mobility trend was calculated over the week prior to the rebound date. (**B**) Subsequent increases in mobility were quantified in terms of daily percent changes relative to the 1-week average prior to the rebound dates shown in figure A. The figure shows the average percent change in the last observation week August 10, 2020 - August 16, 2020 for each age band and each location.

**Figure S2:**
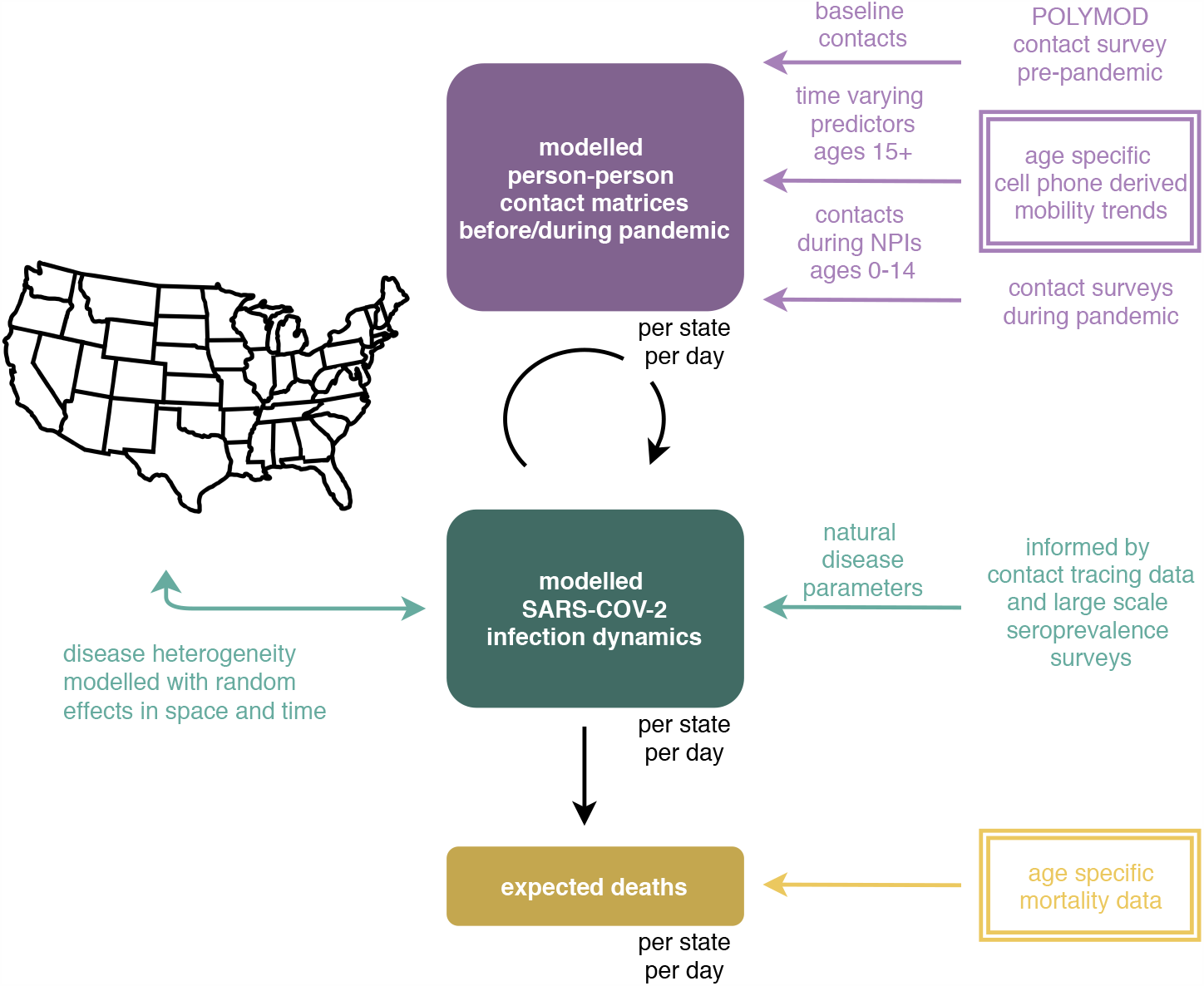
Overview of the age-specific contact and infection model. In the model, SARS-CoV-2 spreads via person-to-person contacts. Person-to-person contacts are described at the population level with the expected number of contacts made by one individual, referred to as contact intensities. Contact intensities are age-specific. Contact intensities vary across locations (states and metropolitan areas) according to each location’s age composition and population density, and change over time. Data from contact surveys before the pandemic are used to define baseline contact intensities. Data from age-specific, cell phone derived mobility trends are used to estimate changes in contact intensities during the epidemic in each location, among individuals aged 15+. Contact intensities involving individuals aged 0-14 are defined based on contact surveys conducted during school closure periods. Infection dynamics in each location are modelled through age-specific, discrete-time renewal equations over time-varying contact intensities. Natural disease parameters such as age-specific susceptibility to infection, the generation time distribution, and symptom onset and onset to death distributions are informed by epidemiologic analyses of contact tracing data. Age-specific infection fatality ratio estimates are informed by large-scale sero-prevalence surveys. Disease heterogeneity is modelled with random effects in space and time on contact intensities and disease parameters. The model returns the expected number of COVID-19 deaths over time in each location, which is fitted against age-specific, COVID-19 mortality data. New data sources presented in this study are indicated in double-framed boxes.

**Figure S3:**
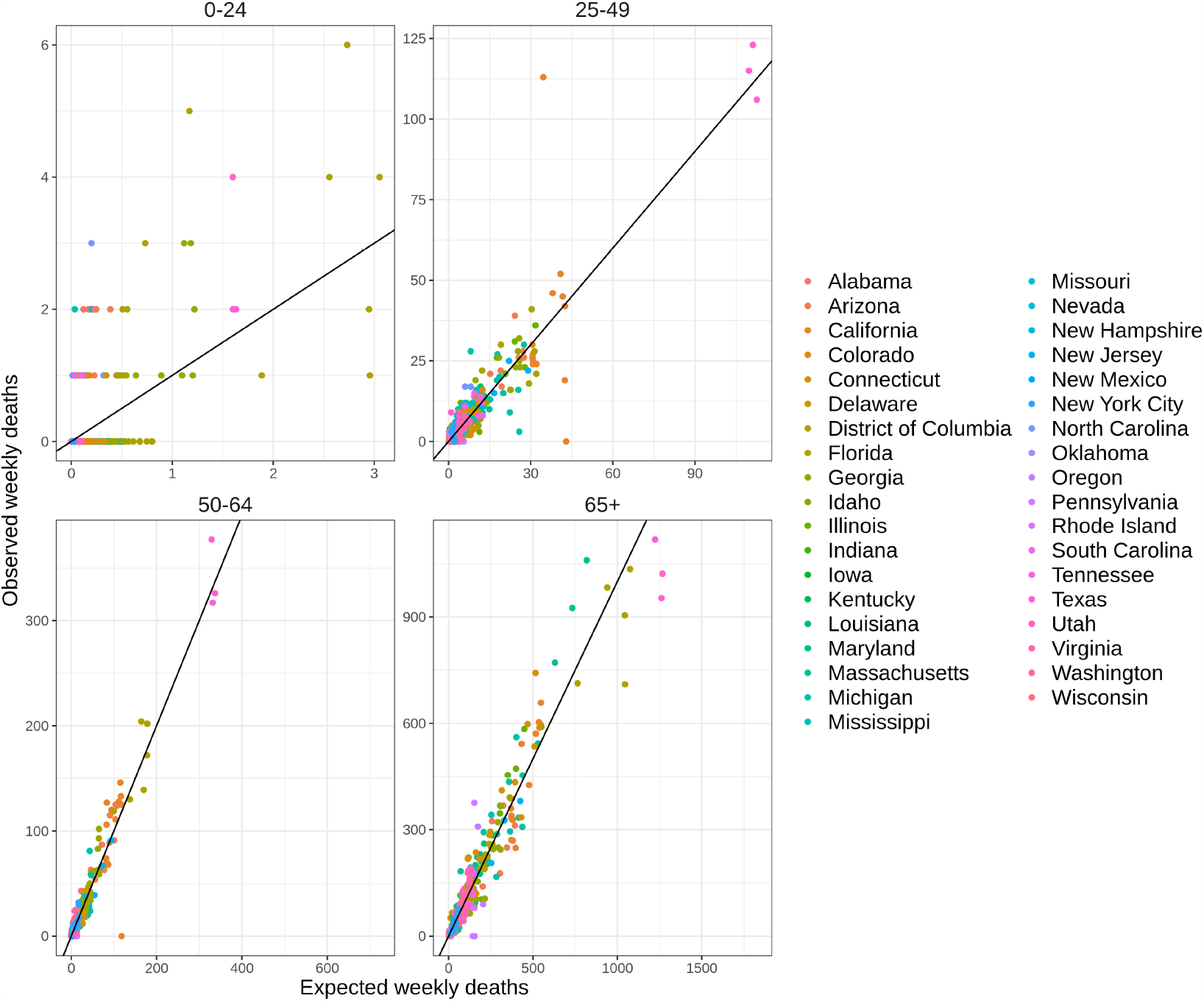
Summary of model fit to age-specific COVID-19 attributable mortality data. To investigate model fit, observed weekly deaths are plotted against posterior median estimates of the expected number of weekly deaths. Locations (states and metropolitan areas) are shown in color. For clarity, the data and weekly estimates are grouped into four age bands.

**Figure S4:**
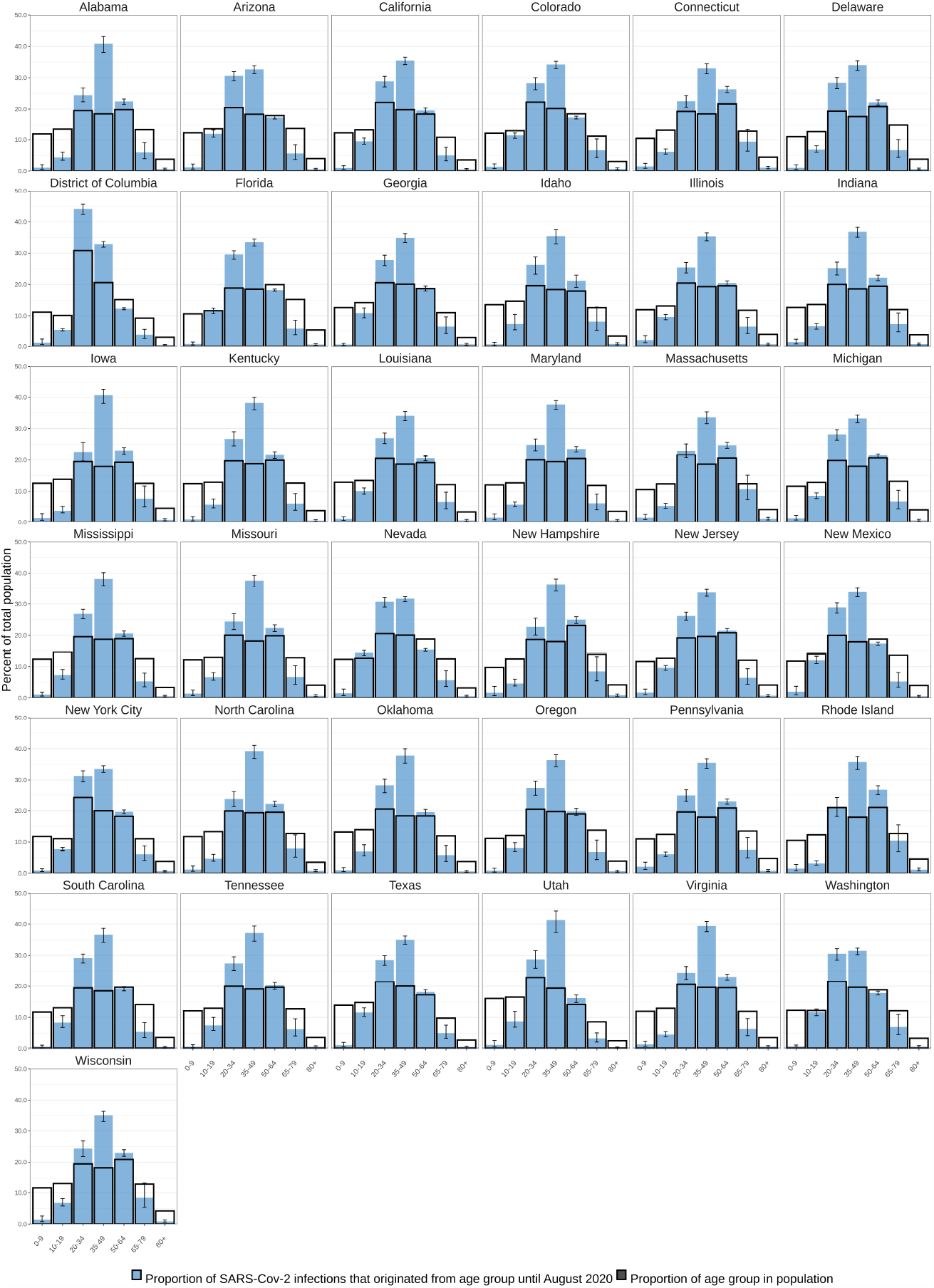
Estimated cumulated contribution of age groups to SARS-CoV-2 infections until August 17, 2020, versus the proportion of the population in the same age group. Posterior median estimates for the percent contributions are shown for each age band (blue) with 95% credible intervals. Age compositions of each state are shown in black.

**Figure S5:**
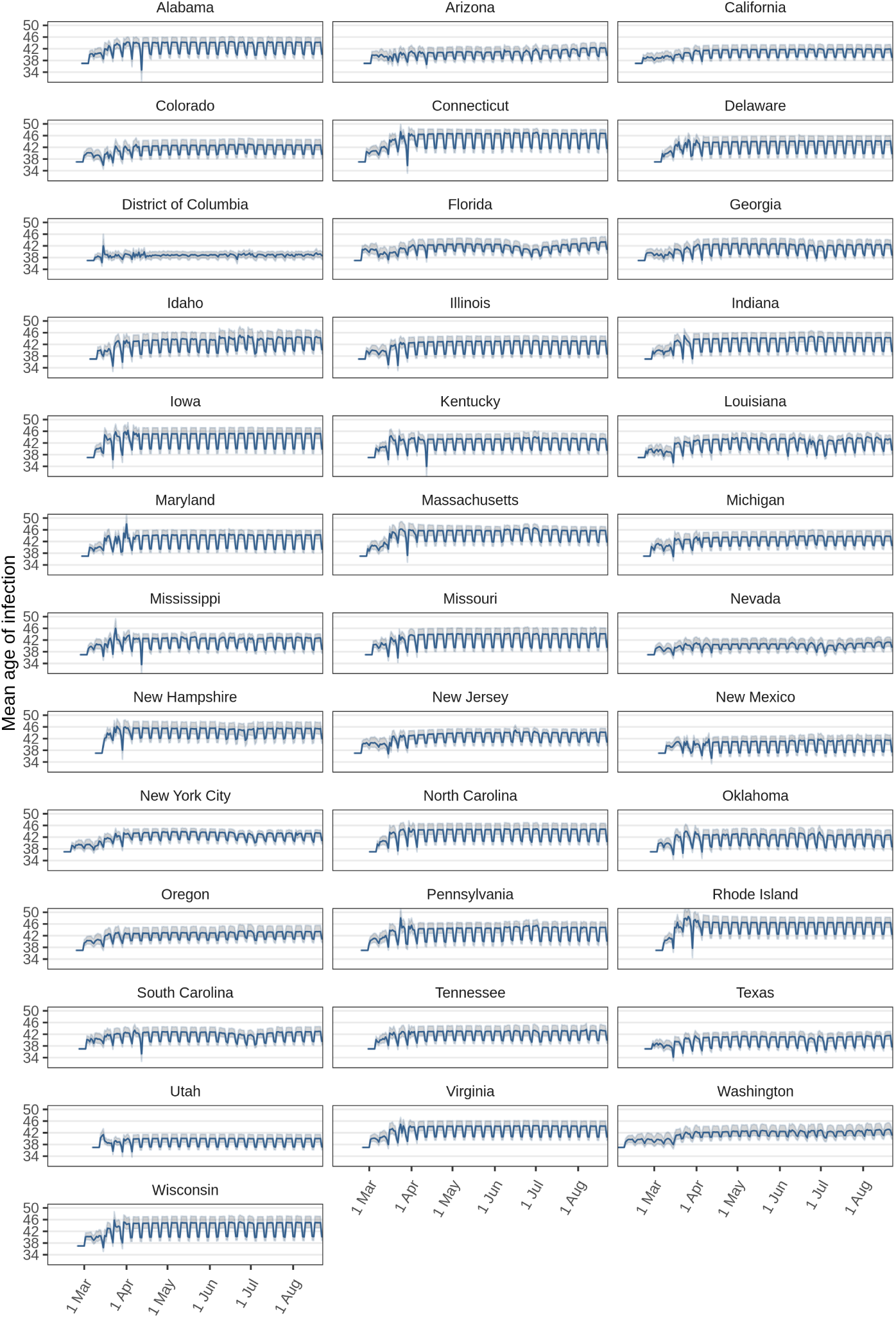
Estimated mean age of SARS-CoV-2 infections. Posterior mean estimates (blue) are shown along with 95% credible intervals (grey). In the first 6 days of reconstructed transmission dynamics, cases were assumed to originate from adults aged 20-54, and trends at the beginning of March reflect a transition from the assumed age composition of initial cases.

**Figure S6:**
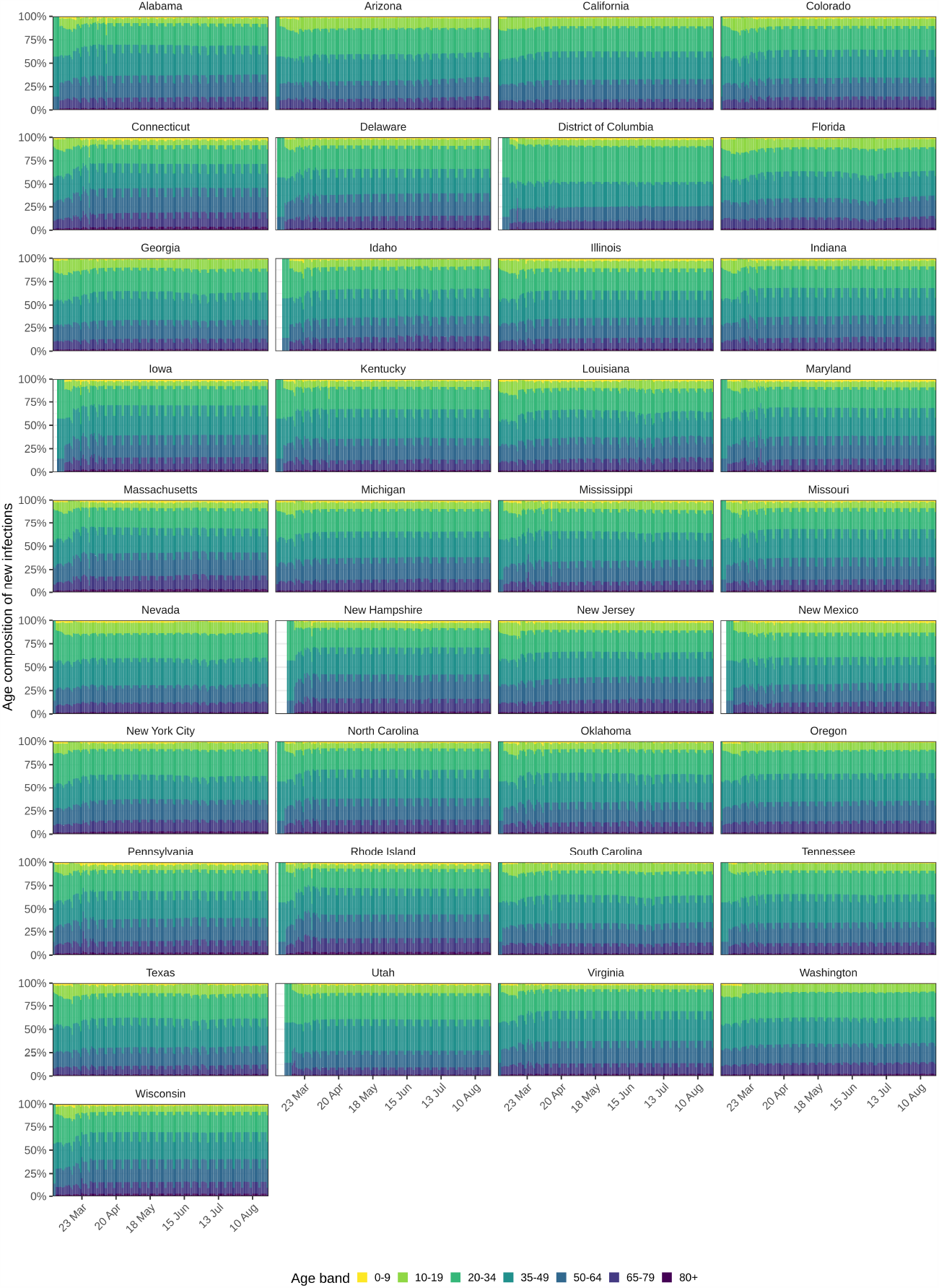
Estimated percent contribution of age groups to SARS-CoV-2 infections. Posterior median estimates are shown for each age band (colours). In the first 6 days of reconstructed transmission dynamics, cases were assumed to originate from adults aged 20-54, and trends at the beginning of March reflect a transition from the assumed age composition of initial cases.

**Figure S7:**
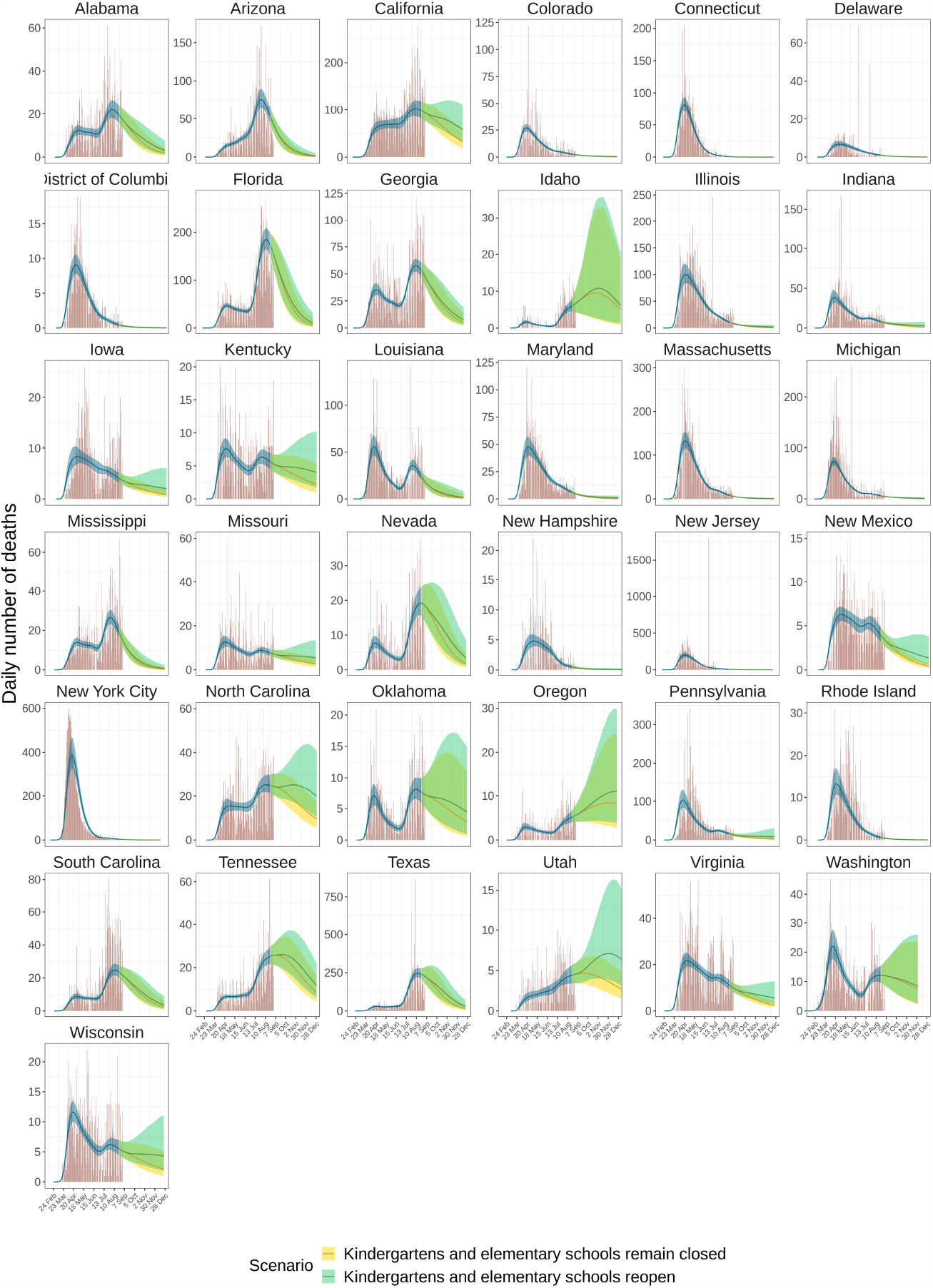
Predicted COVID-19-attributable deaths in the central kindergarten and elementary school reopening scenario. Posterior median estimates (line) are shown along with 95% confidence interval (shaded area). Daily COVID-19-attributable deaths as reported from [2] are overlaid (red bars). Estimated expected deaths are shown in blue for the observation period. Predicted expected deaths in the continued school closure scenario are shown in green. Predicted expected deaths in the school re-opening scenario are shown in yellow. This scenario assumes a 50% transmission reduction from and to children aged 0-11 due to face mask use and other non-pharmaceutical interventions, see Supplementary Text S3.7.

**Figure S8:**
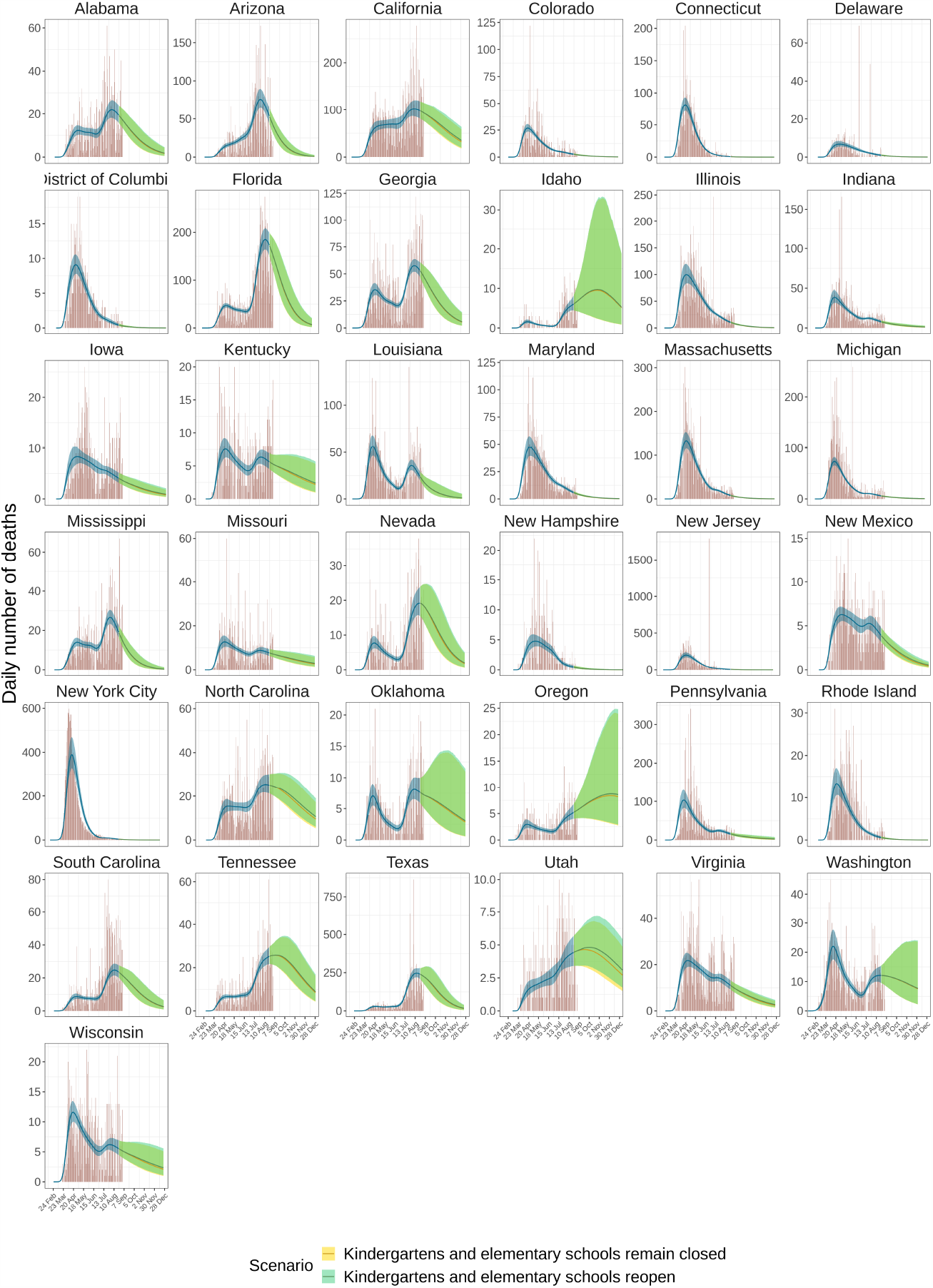
Predicted COVID-19-attributable deaths in the kindergarten and elementary school re-opening scenario with 80% transmission reduction. Posterior median estimates (line) are shown along with 95% confidence interval (shaded area). Daily COVID-19-attributable deaths as reported from [2] are overlaid (red bars). Estimated expected deaths are shown in blue for the observation period. Predicted expected deaths in the continued school closure scenario are shown in green. Predicted expected deaths in the school re-opening scenario are shown in yellow. This scenario assumes a 80% transmission reduction from and to children aged 0-11 due to face mask use and other non-pharmaceutical interventions, see Supplementary Text S3.7.

**Figure S9:**
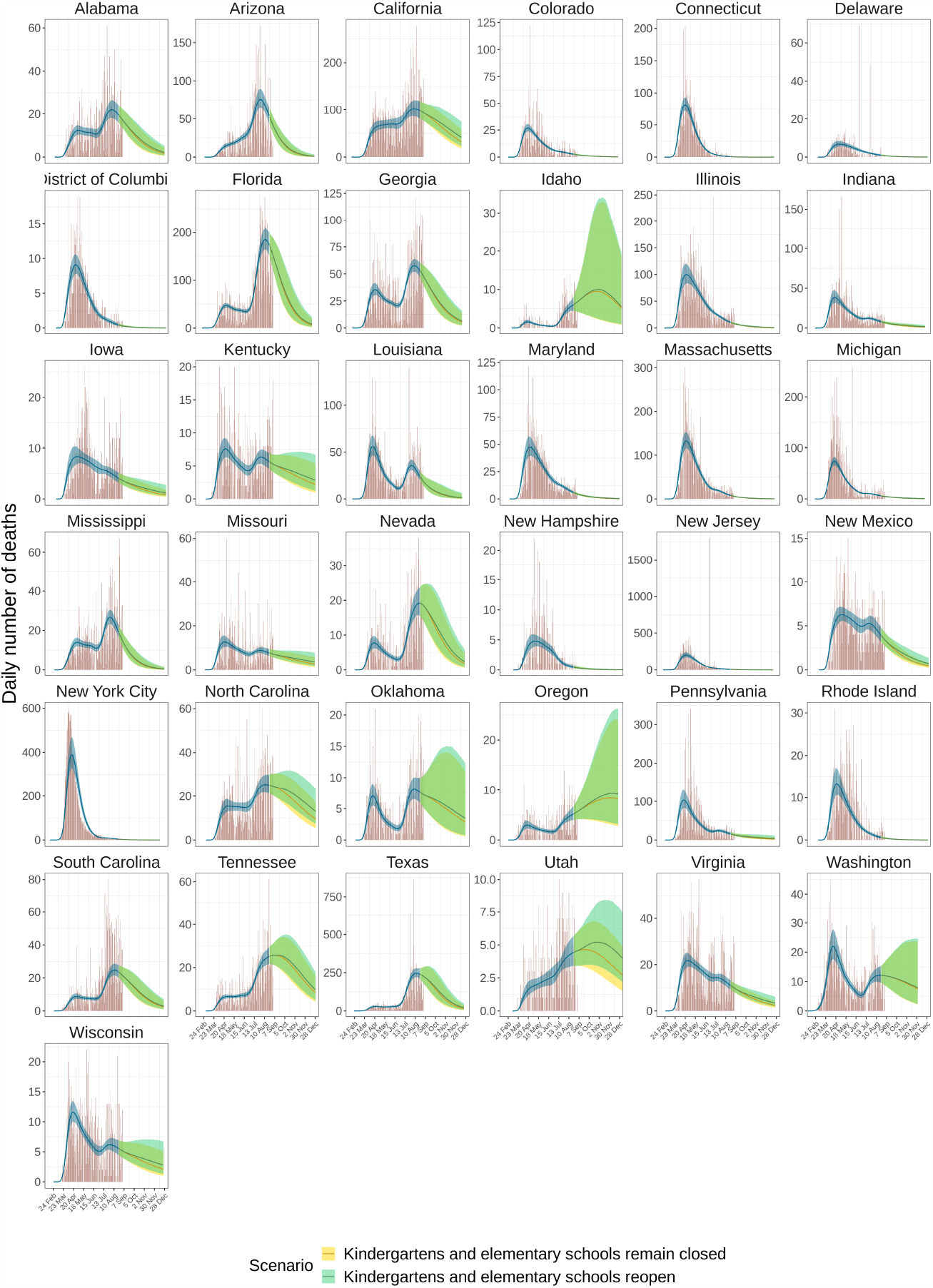
Predicted COVID-19-attributable deaths in the kindergarten and elementary school re-opening scenario with 66% transmission reduction. Posterior median estimates (line) are shown along with 95% confidence interval (shaded area). Daily COVID-19-attributable deaths as reported from [2] are overlaid (red bars). Esttimated expected deaths are shown in blue for the observation period. Predicted expected deaths in the continued school closure scenario are shown in green. Predicted expected deaths in the school re-opening scenario are shown in yellow. This scenario assumes a 66% transmission reduction from and to children aged 0-11 due to face mask use and other non-pharmaceutical interventions, see Supplementary Text S3.7.

**Figure S10:**
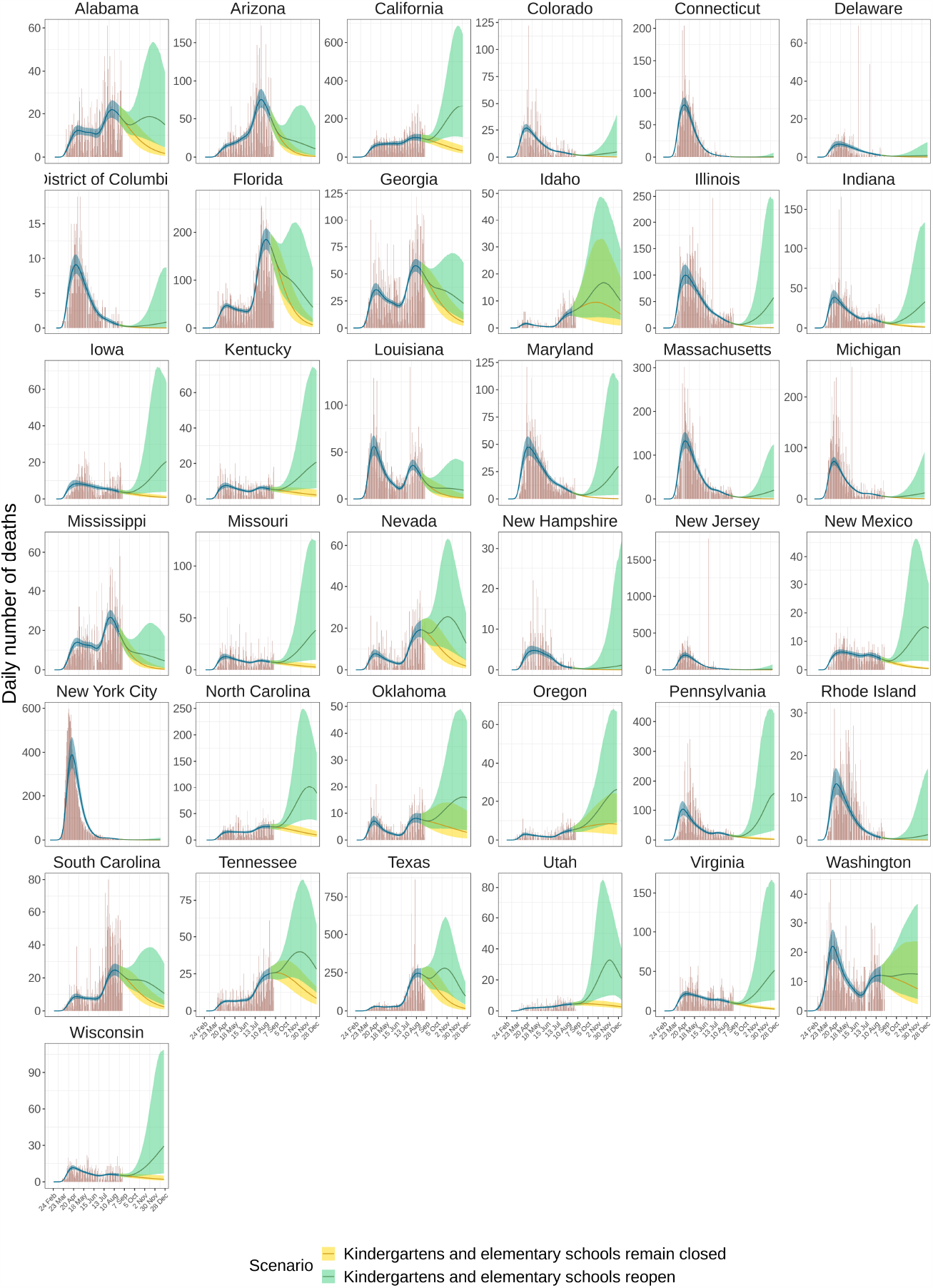
Predicted COVID-19-attributable deaths in the kindergarten and elementary school re-opening scenario with no additional transmission reduction. Posterior median estimates (line) are shown along with 95% confidence interval (shaded area). Daily COVID-19-attributable deaths as reported from [2] are overlaid (red bars). Estimated expected deaths are shown in blue for the observation period. Predicted expected deaths in the continued school closure scenario are shown in green. Predicted expected deaths in the school re-opening scenario are shown in yellow. This scenario assumes no additional transmission reduction from and to children aged 0-11 due to face mask use and other non-pharmaceutical interventions, see Supplementary Text S3.7.

## References

[1] Qun Li et al. “Early transmission dynamics in Wuhan, China, of novel coronavirus–infected pneumonia”. In: New England journal of Medicine (2020).

[2] Johns Hopkins University. “COVID-19 Dashboard”. Available at https://coronavirus.jhu.edu/map.html. 2020.

[3] European Center for Disease Prevention and Control. “Coronavirus disease 2019 (COVID-19) in the EU/EEA and the UK – eleventh update: resurgence of cases”. Available at https://www.ecdc.europa.eu/en/publications-data/rapid-risk-assessment-coronavirus-disease-2019-covid-19-eueea-and-uk-eleventh. 2020.

[4] Florida Division of Emergency Management. “Coronavirus: characteristics of Florida resident cases”. Available at https://www.floridadisaster.org/covid19/covid-19-data-reports/. 2020.

[5] Maine Center for Disease Control & Prevention. “Maine COVID-19 summary”. Available at https://www.maine.gov/dhhs/mecdc/infectious-disease/epi/airborne/coronavirus/data.shtml. 2020.

[6] Joël Mossong et al. “Social contacts and mixing patterns relevant to the spread of infectious diseases”. In: PLoS Med 5.3 (2008), e74.

[7] Thang Hoang et al. “A Systematic Review of Social Contact Surveys to Inform Transmission Models of Close-contact Infections”. In: Epidemiology 30.5 (Sept. 2019), pp. 723–736. URL: https://doi.org/10.1097/ede.0000000000001047.

[8] Juanjuan Zhang et al. “Changes in contact patterns shape the dynamics of the COVID-19 outbreak in China”. In: Science (2020).

[9] Christopher I. Jarvis et al. “Quantifying the impact of physical distance measures on the transmission of COVID-19 in the UK”. In: BMC Medicine 18 (124 2020).

[10] Dennis M. Feehan and Ayesha Mahmud. “Quantifying interpersonal contact in the United States during the spread of COVID-19: ?rst results from the Berkeley Interpersonal Contact Study”. In: medrXiv (2020).

[11] Waksman, A. “Phones, Lambdas and the Joy of Snap-to-Place Technology”. Available at https://enterprise.foursquare.com/intersections/article/phones-lambdas-and-the-joy-of-snap-to-place-techn/. 2018.

[12] Google LLC. “COVID-19 Community Mobility Reports”. Available at https://www.google.com/covid19/mobility. 2020.

[13] J. C. Lee et al. “See How All 50 States Are Reopening (and Closing Again)”. In: (2020). URL: https://www.nytimes.com/interactive/2020/us/states-reopen-map-coronavirus.html.

[14] Nathalie E. Williams et al. “Measures of Human Mobility Using Mobile Phone Records Enhanced with GIS Data”. In: PLOS ONE 10.7 (July 2015), pp. 1–16. URL: https://doi.org/10.1371/journal.pone.0133630.

[15] Roy M Anderson et al. “Epidemiology, transmission dynamics and control of SARS: the 2002–2003 epidemic”. In: Philosophical Transactions of the Royal Society of London. Series B: Biological Sciences 359.1447 (2004), pp. 1091–1105.

[16] Edward Goldstein, Marc Lipsitch, and Muge Cevik. “On the effect of age on the transmission of SARS-CoV-2 in households, schools and the community”. In: medRxiv (2020). URL: https://www.medrxiv.org/content/early/2020/07/28/2020.07.19.20157362.

[17] Nicholas G Davies et al. “Age-dependent effects in the transmission and control of COVID-19 epidemics”. In: Nature Medicine 26 (2020), pp. 1205–1211.

[18] Anita S Iyer et al. “Dynamics and signi?cance of the antibody response to SARS-CoV-2 infection”. In: medRxiv (2020). URL: https://www.medrxiv.org/content/early/2020/07/20/2020.07.18.20155374.

[19] Derek K Chu et al. “Physical distancing, face masks, and eye protection to prevent person-to-person transmission of SARS-CoV-2 and COVID-19: a systematic review and meta-analysis”. In: The Lancet (2020).

[20] Kylie EC Ainslie et al. “Evidence of initial success for China exiting COVID-19 social distancing policy after achieving containment”. In: Wellcome Open Research 5 (2020).

[21] H Unwin et al. “Report 23 - State-level tracking of COVID-19 in the United States: A subnational analysis with future scenarios”. In: Imperial College London COVID-19 reports (2020).

[22] Imperial College London COVID-19 Response Team. “COVID-19 Age specific Mortality Data Repository”. Available at https://github.com/ImperialCollegeLondon/US-covid19-agespecific-mortality-data. 2020.

[23] Meira Levinson, Muge Cevik, and Marc Lipsitch. “Reopening Primary Schools during the Pandemic”. In: New England Journal of Medicine 383 (2020), pp. 981–985.

[24] Enrico Lavezzo et al. “Suppression of a SARS-CoV-2 outbreak in the Italian municipality of Vo’”. In: Nature (2020), pp. 1–5.

[25] Andrew T Levin et al. “Assessing the age specificity and infection fatality rates for COVID-19: systematic review, meta-analysis, and public policy implications”. In: medRxiv (2020). URL: https://www.medrxiv.org/content/early/2020/08/14/2020.07.23.20160895.

[26] Fiona P. Havers et al. “Seroprevalence of Antibodies to SARS-CoV-2 in 10 Sites in the United States, March 23-May 12, 2020”. In: JAMA Internal Medicine (July 2020). URL: https://doi.org/10.1001/jamainternmed.2020.4130.

[27] Centers for Disease Control and Prevention. “Transcript for the CDC Telebriefing Update on COVID-19, June 25, 2020”. In: (2020). URL: https://www.cdc.gov/media/releases/2020/t0625-COVID-19-update.html.

[28] Yang Liu et al. “Association between age and clinical characteristics and outcomes of COVID-19”. In: European Respiratory Journal 55.5 (2020).

[29] W. Messner and S.E. Payson. “Variation in COVID-19 outbreaks at the US state and county levels”. In: Public Health 187 (2020), pp. 15–18. ISSN: 0033-3506.

[30] Qin-Long Jing et al. “Household secondary attack rate of COVID-19 and associated determinants in Guangzhou, China: a retrospective cohort study”. In: The Lancet Infectious Diseases (2020).

[31] European Center of Disease Prevention and Control. “COVID-19 in children and the role of school settings in COVID-19 transmission”. Available at https://www.ecdc.europa.eu/en/publications-data/children-and-school-settings-covid-19-transmission. 2020.

